# Long-term systemic and mucosal SARS-CoV-2 IgA response and its association with persistent smell and taste disorders

**DOI:** 10.1101/2023.01.13.23284341

**Authors:** Jessica Denis, Annabelle Garnier, Laurence Cheutin, Audrey Ferrier, Hawa Timera, Fanny Jarjaval, Carine Hejl, Emmanuelle Billon-Denis, Percy ImmunoCovid group, Damien Ricard, Jean-Nicolas Tournier, Aurélie Trignol, Marie Mura

## Abstract

Current approved COVID-19 vaccines, notably mRNA and adenoviral vectored technologies, still fail to fully protect against infection and transmission of various SARS-CoV-2 variants. The mucosal immunity at the upper respiratory tract represents the first line of defense against respiratory viruses such as SARS-CoV-2 and is thus critical to develop vaccine blocking human-to-human transmission. We measured systemic and mucosal Immunoglobulin A (IgA) response in serum and saliva from 133 healthcare workers from Percy teaching military hospital following a mild infection (SARS-CoV-2 Wuhan strain, n=58) or not infected (n=75), and after SARS-CoV-2 vaccination (Vaxzevria®/Astrazeneca and/or Comirnaty®/Pfizer). While serum anti-SARS-CoV-2 Spike IgA response lasted up to 16 months post-infection, IgA response in saliva had mostly fallen to baseline level at 6 months post-infection. Vaccination could reactivate the mucosal response generated by prior infection, but failed to induce a significant mucosal IgA response by itself. As breakthrough infections have been correlated with IgA levels, other vaccine platforms inducing a better mucosal immunity are needed to control COVID-19 infection in the future. Early post-COVID-19 serum anti-Spike-NTD IgA titer correlated with seroneutralization titers. Interestingly, its saliva counterpart positively correlated with persistent smell and taste disorders more than one year after mild COVID-19, and could potentially be used as an early prognosis biomarker.

## 1. Introduction

Protection against infection and mitigation of human-to-human transmission are key factors to block the spread of respiratory viruses, like influenza virus and SARS-CoV-2 (1, 2). These goals designed by herd immunity have not yet been met by current SARS-CoV-2 approved vaccines (3, 4). Systemic vaccination by mRNA and adenovirus-vectored vaccines has shown a great potential in crisis management but fails to induce a prolonged protection against infection and human-to-human transmission by the subsequent variants (5), even though they could reduce the period of transmission by a faster control of viral replication (6). There is a need for second generation vaccines that may broaden the immune response against multiple variants, induce a long-lasting memory response and protect more efficiently against transmission and breakthrough infections, ideally by imprinting a strong mucosal immune response in the upper respiratory tract, as described in recent preclinical studies (7-9).

The protection of the respiratory tract is particularly complex since it is provided by the integrity of the epithelial barriers, a protective mucus layer propelled by cilia, antimicrobial peptides and innate immune cells as well as by an adaptive immune response initiated at the inductive sites of nasopharynx-associated-lymphoid tissue (NALT) (10). The local microbiota has also been described as a modulator of the immune response (11). Polymeric immunoglobulins A (IgA) are specific soluble mediators of the adaptive immune response. While IgA are monomeric in human serum, they are produced locally mostly under dimeric form in mucosal tissues and are released with the secretory component at the mucosal lumen as secretory IgA (SIgA). Monomeric Ig are also present in mucosal secretions following passive transport from blood compartment, with a majority of IgG and fewer IgA (12). Natural polyreactive IgA with cross-reactivity and low affinity have also been described in the lumen of mucosal surfaces (13). In contrast to nasal or bronchoalveolar samples, saliva is an easy-to-access biofluid where specific SARS-CoV-2 IgA have been detected (14-16), providing information about the mucosal response by harboring a significant population of IgA-secreting plasma cells.

Another major health issue of the COVID-19 pandemic is the persistence of long-lasting clinical disorders (so-called long COVID or post-acute sequelae of COVID-19) for a substantial proportion of SARS-CoV-2-infected people, with heterogeneity due to geographic location, SARS-CoV-2 variants and studied population(17, 18). The clinical presentation is highly diverse, involving several organs and systems. Smell and taste disorders were particularly frequent since the initial phase of the COVID-19 pandemic (19, 20), with quantitative (hypo-, hyper-, anosmia or ageusia) or qualitative (dys-, phantosmia or phantageusia) alterations. Persistent olfactory disorders have been notified from 10% to more than 50% according to SARS-CoV-2 variants (18, 19), and can be recovered in a few months. Nevertheless, recent data described less than 40% people with a complete recovery after 2 years, and 7.5% displaying no improvement (21). The pathophysiology of long COVID remains poorly understood and may include viral persistence or delayed clearance, autoimmunity and tissue damages due to inflammation. Recent studies have also highlighted the persistence of the humoral response (22, 23). The contribution of the mucosal humoral compartment has started to be investigated to potentially identify prognosis markers (24).

In this study, we therefore examined the anti-SARS-CoV-2 IgA response over more than one-year in serum and saliva from a cohort of 133 healthcare workers suffering from a mild COVID-19 (SARS-CoV-2 Wuhan strain, n=58, the COVID+ group), and non-infected controls (n=75, the COVID-group). We analyzed the antigen specificity of the response against the spike immune-dominant target and its subdomains (Spike (S), Spike receptor binding domain (RBD), Spike N terminal domain (NTD)) and the nucleoprotein (N). We evaluated the impact of pre-exposure to other human coronaviruses (HCoV-NL63, HCoV-229E, HCoV-OC43, HCoV-HKU1) and the reactivation of mucosal immunity after systemic intramuscular vaccination. Post-infection anti-SARS-CoV-2 Spike IgA response in serum was sustained up to 16 months, whereas IgA response in saliva was back to its baseline level after 6 months. We observed that vaccination reactivated prior mucosal immune response generated by infection, while vaccine alone failed to induce significant IgA titers in saliva. Finally, the initial levels of serum anti-Spike N-terminal domain (NTD) IgA were found to positively correlate with seroneutralization efficacy, whereas their salivary counterpart positively correlated with the persistence of smell disorders or taste disorders more than one year after acute COVID-19.

## 2. Results

### 2.1. Serum anti-SARS-CoV-2 IgA were maintained for up to 16 months post-infection and NTD-specificity positively correlated with seroneutralization titers

The main characteristics of the cohort are represented in Table 1. In the total of 400 volunteers achieving the totality of the longitudinal follow-up, we selected 133 individuals (75 naive individuals and 58 previously-infected individuals), based on the certainty of the diagnosis at enrollment, the absence of a new COVID-19 infection during the longitudinal follow-up, and the quality/quantity of saliva samples (Supplementary method and supplementary figure S1).

**Table 1:**
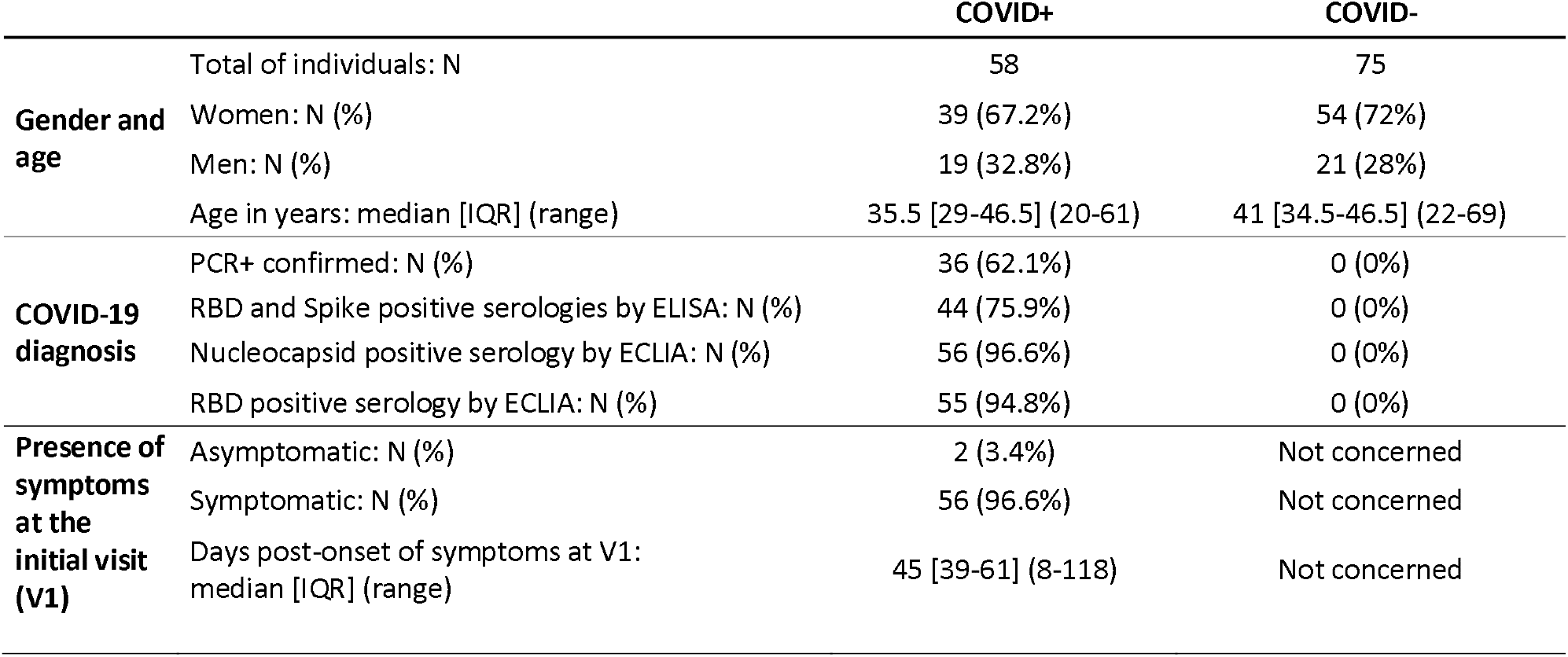
Main characteristics of naive individuals (COVID-) and previously infected individuals (COVID+) at the initial visit (V1).

At enrollment (Visit 1 – V1), the median onset of symptoms was 1.5 months (Table 1) and specific anti-SARS-CoV-2 Spike IgA were present in the serum from almost all previously infected individuals (n=56/58) (Figure 1A). The positive threshold of each test was determined by the Youden index (ROC curves in supplementary figure S2). The IgA antibodies targeted preferentially the Spike/RBD (n=54/58) (Figure 1B) than the Spike/NTD (n=48/58) (Figure 1C), and the nucleocapsid-N (n=48/58) (Figure 1D). Six months later (Visit 2 – V2), the IgA signal significantly decreased, regardless of the targeted antigen, as compared to V1 (p<0,001) (Figure 1). Especially, the mean titer of anti-N IgA fell under the positive threshold at 6 months. Hypothesizing a linear evolution of the log(IgA signal) over time between V1 and V2, the modeling using linear mixed models found negative slopes (Supplementary Table S1) that corresponded to a daily decrease of IgA concentration of 2.70 ‰ for S, 2.78 ‰ for Spike/RBD, 3.95 ‰ for Spike/NTD and 6.52 ‰ for N. Between 6 and 12 months following the inclusion (V2 and V3 respectively), anti-N IgA had a tendency to decrease (Figure 1D), with a negative modeling slope corresponding to a daily decrease of 0.21 ‰, whereas we observed a stabilization of IgA titers against all other targets. The general anti-SARS-CoV-2 serological response (anti-N and anti-Spike/RBD total Ig and seroneutralization titers) from previously infected individuals is shown in figure 2. The Spike/RBD total Ig response (Figure 2A) increased over time (p<0.001), whereas the anti-N total Ig response (Figure 2B) decreased (p<0.001). The seroneutralization capacity of the serum against the BetaCoV/France/IDF0372/2020 strain was maintained up to 16 months post-infection (Figure 2C).

**Figure 1:**
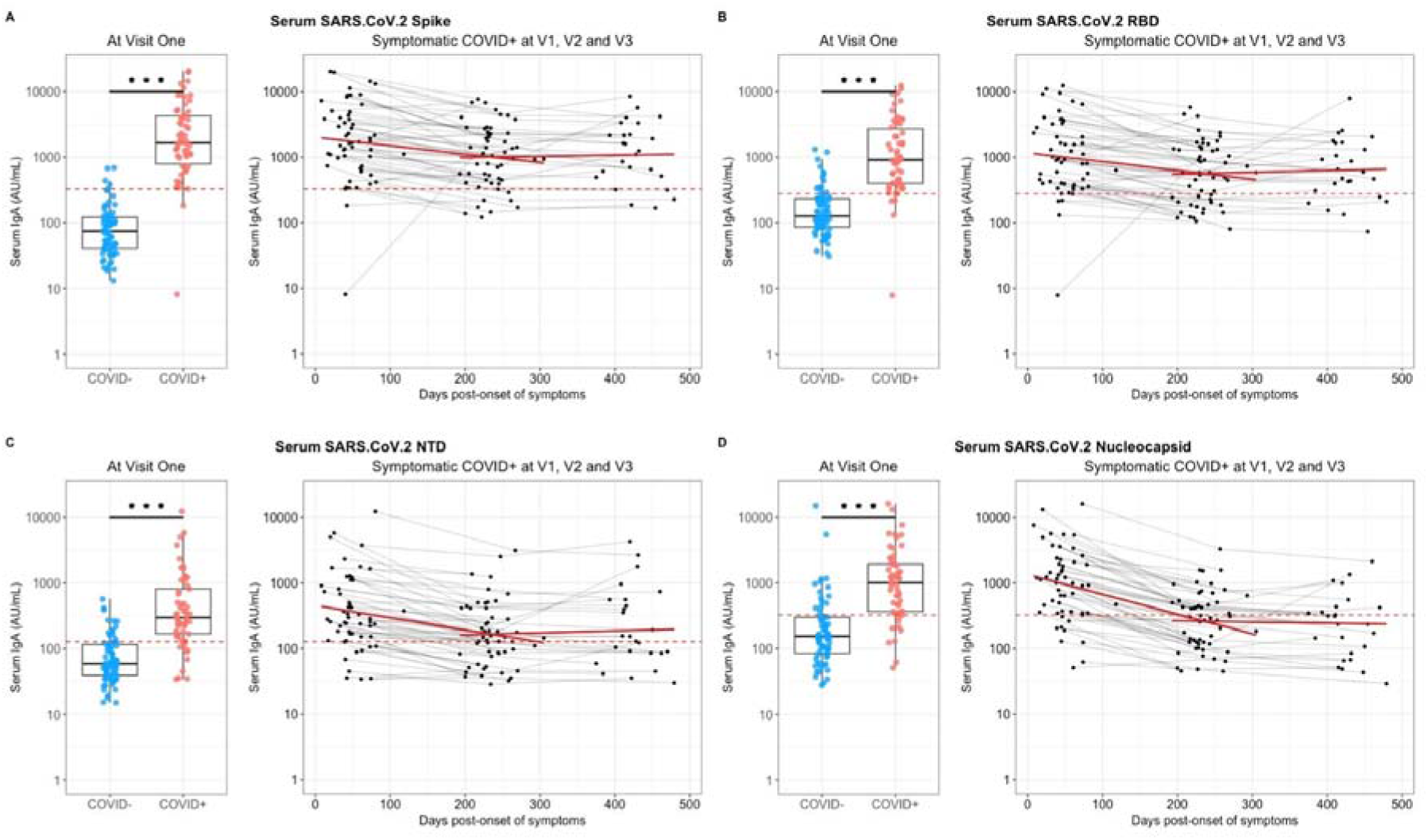
Serum anti-SARS-CoV-2 IgA kinetics and specificity up to 16 months after COVID-19 acute infection and without immunization. Serology titers of anti-SARS-CoV2 IgA (UA/ml) from naive (COVID-, blue) and previously infected (COVID+, red) individuals at V1, and its kinetic of decrease (mean slope in red) for symptomatic COVID-19+ individuals up to 16 months after the acute infection. (A) Serology IgA titers against the whole Spike. (B) Serology IgA titers against the Spike/RBD. (C) Serology IgA titers against the Spike/NTD. (D) Serology IgA titers against the Nucleocapsid. The red dashed line corresponded to the positivity threshold. Wilcoxon-rank sum test: ns = not significant, *** p<0.001. Linear mixed models were used to calculate the mean slope between each visit (red line).

**Figure 2:**
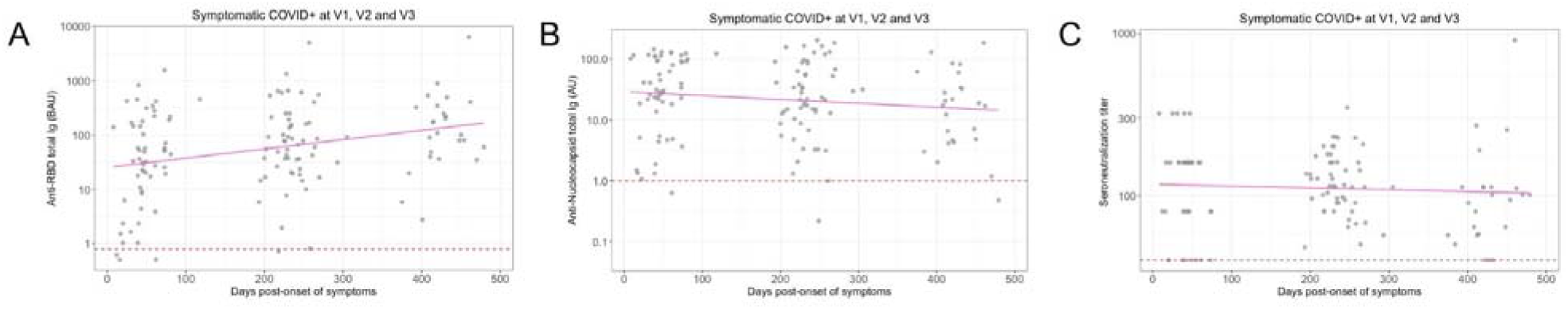
Serum anti-SARS-CoV-2 total Immunoglobulins (Ig) and seroneutralization titers at each visit and without immunization for previously infected individuals. (A) Titers of total Ig targeting the Spike/RBD and (B) the Nucleocapsid in serum from previously infected individuals are represented at enrollment (V1), 6 months (V2) and 12 months (V3) later. (C) Seroneutralization titers from previously infected individuals at V1, V2 and V3. The positivity thresholds are represented with a red dashed line. Linear regression slopes from (A) and (B) were significantly different from zero with p < 0.001.

Seroneutralization titers at V1 positively correlated with serum anti-Spike/NTD IgA (Spearman, rho=0.32, p=0.012) (Supplementary figure S3A). More than 6 months post-infection (V2), seroneutralization titers correlated with serum anti-Spike/NTD IgA (Spearman, rho=0.5, p=0.013) and anti-Spike IgA (Spearman, rho=0.32, p=0.03) (Supplementary figure S3B). More than one year after COVID-19 infection (V3) and without any vaccination, seroneutralization titers still correlated with serum anti-Spike IgA (Spearman, rho=0.3, p=0.02) (Supplementary figure S3C).

Overall, we observed in our cohort an induction of high serological titers of anti-SARS-CoV-2 Spike IgA that positively correlated with seroneutralization titers, and maintained for up to 16 months post-infection despite a first decline after 6 months.

### 2.2. COVID-19 induced weak but significant salivary anti-Spike IgA, followed by a constant decrease over time

At enrollment (V1), specific anti-SARS-CoV-2 IgA were also detected in saliva from infected individuals, as compared to controls individuals (Figure 3). These antibodies targeted the total Spike (p<0.001, figure 3A), preferentially Spike/RBD (p=0.002, figure 3B) than Spike/NTD (p=0.001, figure 3C), but not the nucleocapsid (p=0.43, figure 3D). During the first six months, the IgA level in saliva significantly decreased against all targets (p<0,04). The modelling using linear mixed models found negative slopes that corresponded to a daily decrease of IgA concentration of 1.16 ‰ for S, 0.89 ‰ for Spike/RBD and 0.90 ‰ for Spike/NTD. Between 6 and 12 months, a similar decrease was observed against these targets (supplementary Table S1).

**Figure 3:**
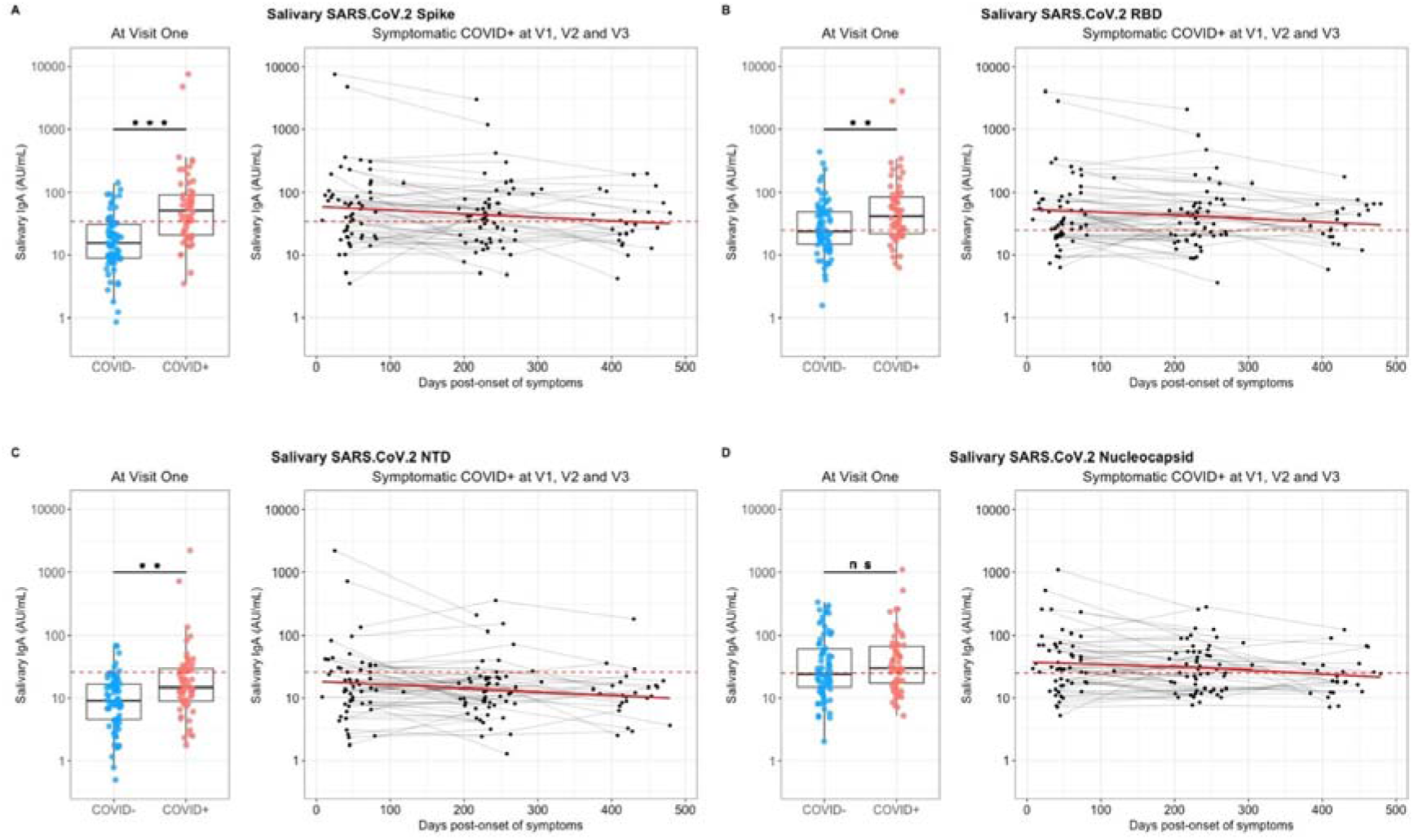
Saliva anti-SARS-CoV-2 IgA kinetics and specificity up to 16 months after COVID-19 acute infection and without immunization. Salivary titers of anti-SARS-CoV2 IgA (UA/ml) from naive (COVID-, blue) and previously infected (COVID+, red) individuals at enrollment (V1), and its kinetic of decrease (mean slope in red) for symptomatic infected individuals up to 16 months after the acute infection. (A) Salivary IgA titers against the whole Spike. (B) Salivary IgA titers against the Spike/RBD. (C) Salivary IgA titers against the Spike/NTD. (D) Salivary IgA titers against the Nucleocapsid. Wilcoxon-rank sum test: ns = not significant, ** p<0.01, *** p<0.001. Linear mixed models were used to calculate the mean slope between each visit (red line).

So, a specific anti-SARS-CoV-2 Spike salivary IgA response was noted shortly after infection, but weak, variable and with a continuous decrease over the time. This was in contrast with anti-N antibodies for which the detected signal did not differ between naive and previously infected individuals.

### 2.3. Post-COVID19 anti-SARS-CoV-2 Spike IgA titers reached other human coronaviruses range in saliva and serum

In order to evaluate the impact of human coronaviruses pre-exposure on the level of anti-SARS-CoV-2 response and estimate the cross-reactivity *in vitro*, we quantified IgA in serum and saliva against the Spike protein from two alpha-coronaviruses (hCoV-229E, hCoV-NL63) and three beta-coronaviruses (hCoV-OC43, hCoV-HKU1, SARS-CoV-1) at V1. No previous exposure to SARS-CoV-1 was reported by any participant of the cohort. First, we observed in the COVID-19-negative group a similar range of detected signal against the four hCoV, with slightly higher responses against hCoV-OC43 and hCoV-229E compared to hCoV-NL63 and hCoV-HKU1 in serum and saliva (Figure 4A). IgA directed against SARS-CoV-1 or SARS-CoV-2 were weakly detected, which was obvious in the absence of exposure to the viruses in this group and may reflect non-specific response/background noise, or low level of *in vitro* cross-reactivity between human coronaviruses.

**Figure 4:**
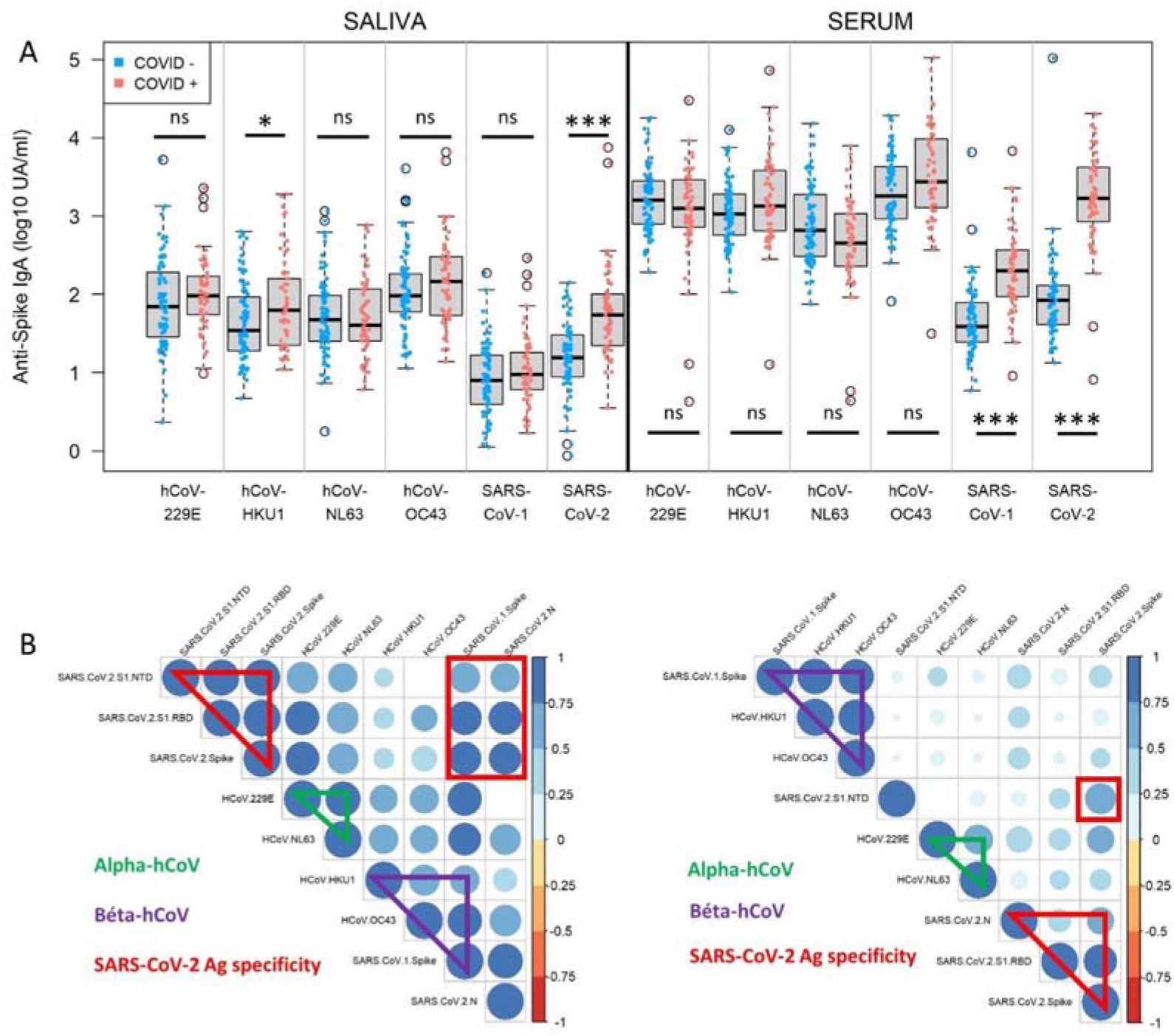
Serum and saliva anti-Spike IgA titers against human coronaviruses. (A) Normalized log-10 transformed serology titers of anti-Spike IgA (log10 UA/ml) in saliva (left) and serum (right) from naive (COVID-, blue) and previously infected (COVID+, red) individuals measured by MSD technology at enrollment (V1). IgA specificity corresponds to the Spike proteins from human coronaviruses (hCoV) 229E, HKU1, NL63, OC43, SARS-CoV-1 and SARS-CoV-2. Kruskal-Wallis test: ns=not significant; * p<0.05 ; *** p<0.001. (B) Correlation matrices in saliva (left) and serum (right) at V1 showing significant positive correlations between alpha-coronaviruses (hCoV 229E and NL63), beta-coronaviruses (hCoV OC43, HKU1, SARS-CoV-1, SARS-CoV-2), as well as SARS-CoV-2 antigen specificity (N, Spike, Spike/RBD, Spike/NTD) in previously infected individuals. Only significant correlations (Spearman, p<0.05) are represented on the matrices. The color and size of the dots (scale next to the graph) indicate the degree of correlation (Spearman, rho) between the different parameters (small to large indicating low to high correlation).

At V1, there were significantly higher signals against hCoV-HKU1 in the saliva (p=0.04) and SARS-CoV-1 in the serum (p<0.001) in the COVID-19+ group than in the COVID-19-negative group. The rest of the signals were unchanged by the COVID-19 status. As these viruses are two beta-coronaviruses belonging to a closer specie, it may be explained by some cross-reactivity *in vitro*. The detected signal against SARS-CoV-2 Spike after infection in this group reached the same level of response observed against the other hCoV in serum and saliva. Then we looked for correlations between individual responses and noticed that in serum (Figure 4B, right panel, Spearman; supplementary figure S4, Pearson) beta-coronaviruses responses correlated together, as well as alpha-coronaviruses, independently of SARS-CoV-2 response. In saliva, the pattern showed a larger correlation between coronaviruses genera (Figure 4B, left panel, Spearman; supplementary figure S5, Pearson).

Thus, we identified some cross-reactivity *in vitro* of anti-SARS-CoV-2 Spike IgA against different human coronaviruses. But this phenomenon did not interfere with specific SARS-CoV-2 IgA response that reached the same range of hCoV IgA titers after infection.

### 2.4. Vaccination reactivated mucosal immunity in previously infected individuals but was not able to induce significant mucosal response in naive individuals

At 12 months of the follow-up, the vaccination campaign had already started for healthcare workers, which were a priority population. The vaccine campaign started at the end of December 2020 and a high proportion of the cohort already got vaccinated. At V3, individuals ranged from 0 to 2 doses of vaccine (Table 2) due to limited supplies, updating of vaccine recommendations and vaccine hesitancy. Two vaccination schedules were conducted at this time (Table 3), with 54 individuals having received first an mRNA vaccine (Pfizer-BioNTech, Comirnaty®) and 32 individuals having received first a ChAd-vectored vaccine (Astrazeneca, Vaxzevria®). Few people (n=14) received a heterologous prime-boost with Vaxzevria® followed by Comirnaty® 12 weeks apart (as compared to 4 weeks apart for 2 doses of Comirnaty®). The remaining vaccinated individuals got two doses of Comirnaty® (n=37). At V3 and without vaccination, previously infected individuals did not have any more detectable circulating anti-N IgA whereas anti-Spike (including Spike/RBD and Spike/NTD) IgA were still present. Both vaccination schedule induced significant titers of anti-SARS-CoV-2 Spike IgA in serum (Supplementary Figure S6), in naive and in previously infected individuals. The different group comparisons with the level of significance are represented in Figure 5. Not surprisingly, the specificity of the response was restricted to the Spike, including Spike/RBD and Spike/NTD, and did not extend to the nucleocapsid, an antigen absent from all vaccines. Nevertheless, even after 2 doses, IgA levels in the serum from COVID-19-negative individuals did not reach the level of response observed in previously infected individuals after one or two doses (p<0.001).

**Table 2:** Vaccination status of naive individuals (COVID-) and previously infected individuals (COVID+) at the last visit (V3).

|  |  | COVID+ | COVID- |
| --- | --- | --- | --- |
| **Participants at visit three (V3)** | Total of individuals: N | 58 | 75 |
| **No vaccine** | N (% of total individuals) | 29 (50%) | 18 (24%) |
|  | Women: N (%) | 20 (69%) | 14 (77.8%) |
|  | Men: N (%) | 9 (31%) | 4 (22.2%) |
|  | Age in years: median [IQR] (range) | 37 [28-47] (20-61) | 37.5 [31.8-42.5] (22-61) |
| **One vaccine dose** | N (% of total individuals) | 20 (34.5%) | 15 (20%) |
|  | Women: N (%) | 14 (70%) | 12 (80%) |
|  | Men: N (%) | 6 (30%) | 3 (20%) |
|  | Age in years: median [IQR] (range) | 34 [29-38.5] (26-55) | 42 [34.5-44] (22-47) |
|  | Days post first vaccine at V3: median [IQR] (range) | 71 [47-98] (10-148) | 93[68-115] (9-140) |
| **Two vaccine doses** | N (% of total individuals) | 9 (15.5%) | 42 (56%) |
|  | Women: N (%) | 5 (55.6%) | 28 (66.7%) |
|  | Men: N (%) | 4 (44.4%) | 14 (33.3%) |
|  | Age in years: median [IQR] (range) | 41 [32-58] (26-59) | 43 [37.3-48.8] (22-69) |
|  | Days post second vaccine at V3: median [IQR] (range) | 86 [19-104] (7-120) | 57 [28-92] (1-113) |
|  | Interval (in days) between 2 doses: median [IQR] (range) | 28 [28-74] (21-84) | 28 [28-47.8] (21-93) |

**Table 3:** Vaccine schedule of naive (COVID-) and previously infected individuals (COVID+) at the last visit (V3).

| Vaccine schedule |  | COVID+ | COVID- |
| --- | --- | --- | --- |
| **One dose: Pfizer** | N (%) | 13 (22.4%) | 4 (5.3%) |
|  | Women: N (%) | 8 (61.5%) | 3 (75%) |
|  | Men: N (%) | 5 (38.5%) | 1 (25%) |
|  | Age in years: median [IQR] (range) | 35 [29-38] (25-55) | 43.5 [38-44.3] (23-45) |
|  | Days post first dose: median [IQR] (range) | 47 [21- 62] (10-111) | 29 [19.5- 44.5] (9-73) |
| **One dose: AstraZeneca** | N (%) | 7 (12.1%) | 11 (14.7%) |
|  | Women: N (%) | 6 (85.7%) | 9 (81.8%) |
|  | Men: N (%) | 1 (14.3%) | 2 (18.2%) |
|  | Age in years: median [IQR] (range) | 33 [30- 38.5] (28-43) | 42 [34.5- 43] (22-47) |
|  | Days post first dose: median [IQR] (range) | 60 [56.5-71] (42-80) | 77 [67.5-81] (61-90) |
| **Two doses: Pfizer/Pfizer** | N (%) | 6 (10.3%) | 31 (41.3%) |
|  | Women: N (%) | 3 (50%) | 18 (58.1%) |
|  | Men: N (%) | 3 (50%) | 13 (41.9%) |
|  | Age in years: median [IQR] (range) | 54.5 [36.8-58] (28-59) | 45 [40-50.5] (22-69) |
|  | Days post second dose: median [IQR] (range) | 98 [87.5-106.3] (77-120) | 87 [44-95.5] (1-113) |
|  | Interval (in days) between 2 doses: median [IQR] (range) | 28 [27.3-28] (21-28) | 28 [27-28] (21-49) |
| **Two doses: AstraZeneca/Pfizer** | N (%) | 3 (5.2%) | 11 (14.7%) |
|  | Women: N (%) | 2 (66.7%) | 10 (90.9%) |
|  | Men: N (%) | 1 (33.3%) | 1 (9.1%) |
|  | Age in years: median [IQR] (range) | 36 [31-38.5] (26-41) | 36 [29-38] (28-58) |
|  | Days post second dose: median [IQR] (range) | 13 [10-16] (7-19) | 19 [6-28] (1-63) |
|  | Interval (in days) between 2 doses: median [IQR] (range) | 79 [76.5-81.5] (74-84) | 77 [68-81] (35-93) |

**Figure 5:**
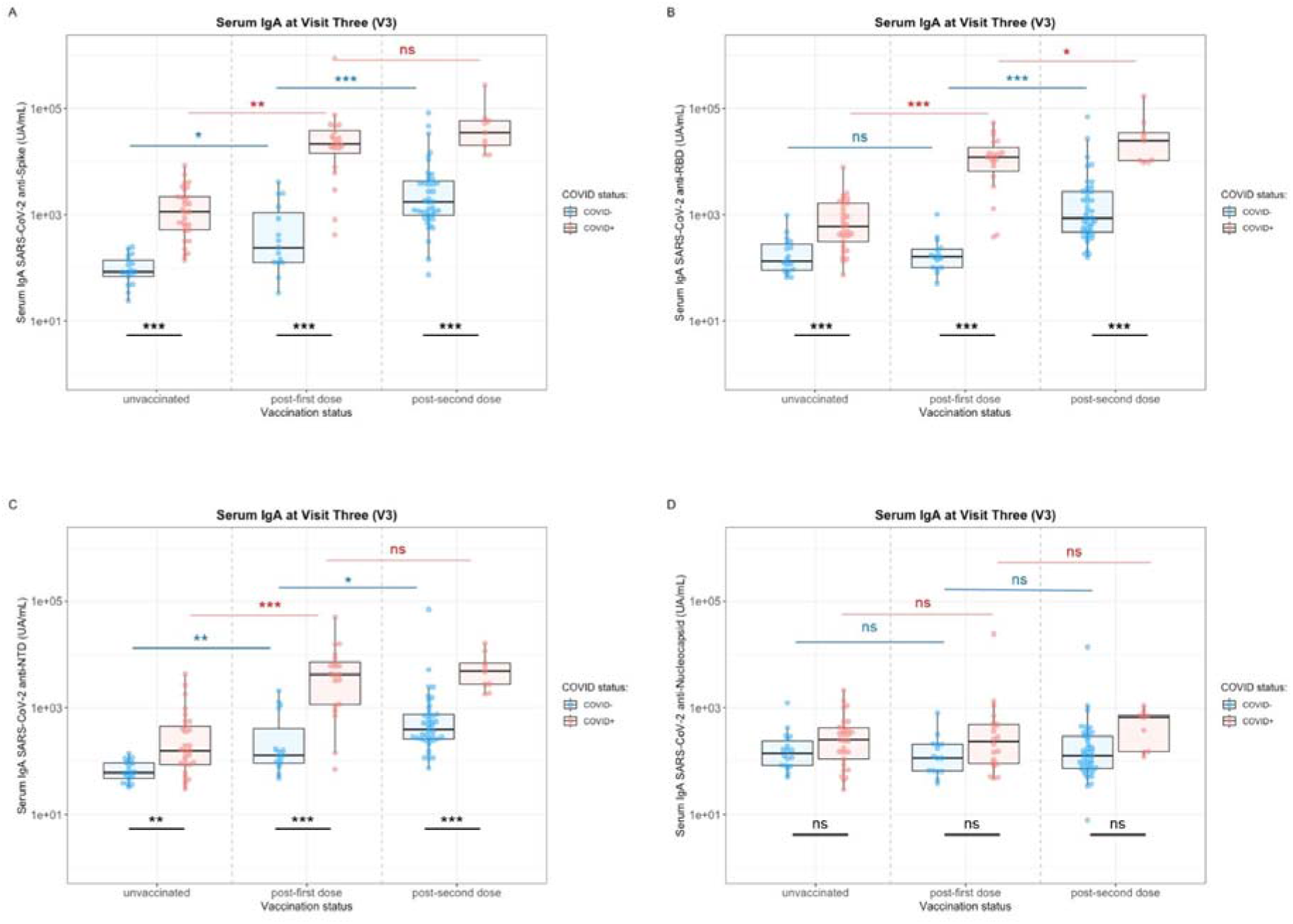
Impact of the number of vaccine doses on serum anti-SARS-CoV-2 IgA titers in the presence (red) or absence (blue) of a previous COVID-19 infection. (A) Serology IgA titers against the whole Spike. (B) Serology IgA titers against the Spike/RBD. (C) Serology IgA titers against the Spike/NTD. (D) Serology IgA titers against the Nucleocapsid. Two-Way ANOVA tests with Tukey post’hoc test: ns = not significant; * p<0.05 ; ** p<0.001; *** p<0.001.

In saliva, we observed significantly higher titers of anti-Spike IgA in individuals previously infected by COVID-19 after one dose (p=0.02) or two doses (p=0.003) of vaccine (Figure 6A). It seemed to have no difference between both vaccination schedules (Supplementary figure S7), when taking into account the variability of the delay between the last vaccine dose and the visit, but we did not have a sufficient number of subjects to statistically confirm it. Conversely, the vaccine alone in naive individuals (one or two doses) did not induce specific salivary anti-Spike IgA. Similar results were observed for SARS-CoV-2 Spike/RBD (Figure 6B) and Spike/NTD-specific IgA (Figure 6C). The anti-Spike IgA signals in the COVID-19-positive group after vaccination (one dose, V3) were highly correlated with the V1 post-COVID-19 response (Adj R^2^=0.735, p<0.001) (Figure 7A). Moreover, the slope of the linear regression between serum and saliva after immunization (one or two doses) was highly different in naive (Adj R^2^=0.31, slope = 1, p<0.001) and previously infected individuals (Adj R^2^=0.44, slope = 0.45, p<0.001) (Figure 7B). These analyses were not affected by the removal of two individuals with extremely high IgA levels in saliva and serum (data not shown), and supported the hypothesis that immunization reactivated previously-induced mucosal immunity.

**Figure 6:**
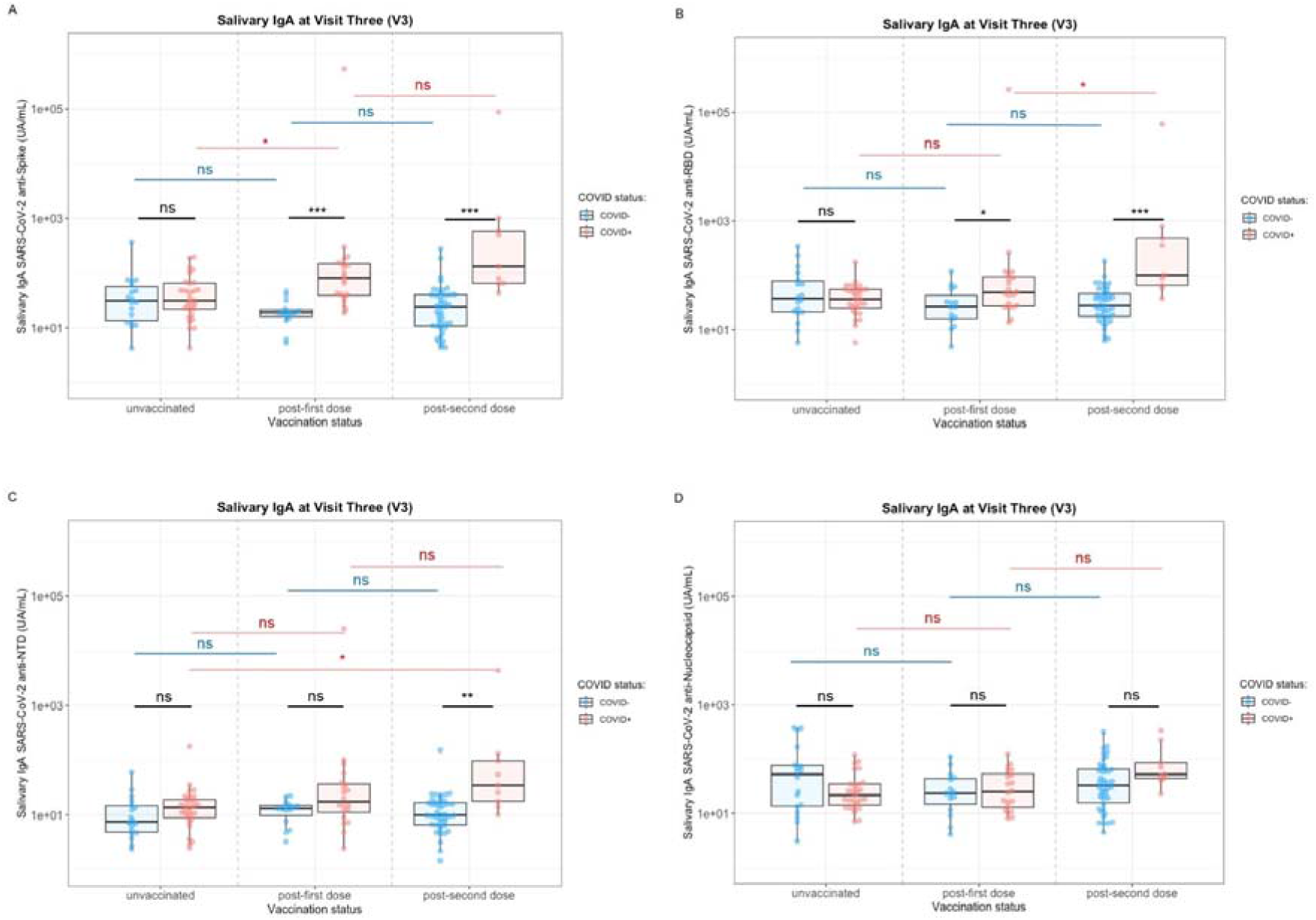
Impact of the number of vaccine doses on saliva anti-SARS-CoV-2 IgA titers in the presence (red) or absence (blue) of a previous COVID-19 infection. (A) Salivary IgA titers against the whole Spike. (B) Salivary IgA titers against the Spike/RBD. (C) Salivary IgA titers against the Spike/NTD. (D) Salivary IgA titers against the Nucleocapsid. Two-Way ANOVA tests with Tukey post’hoc test: ns = not significant; * p<0.05 ; ** p<0.001; *** p<0.001.

**Figure 7:**
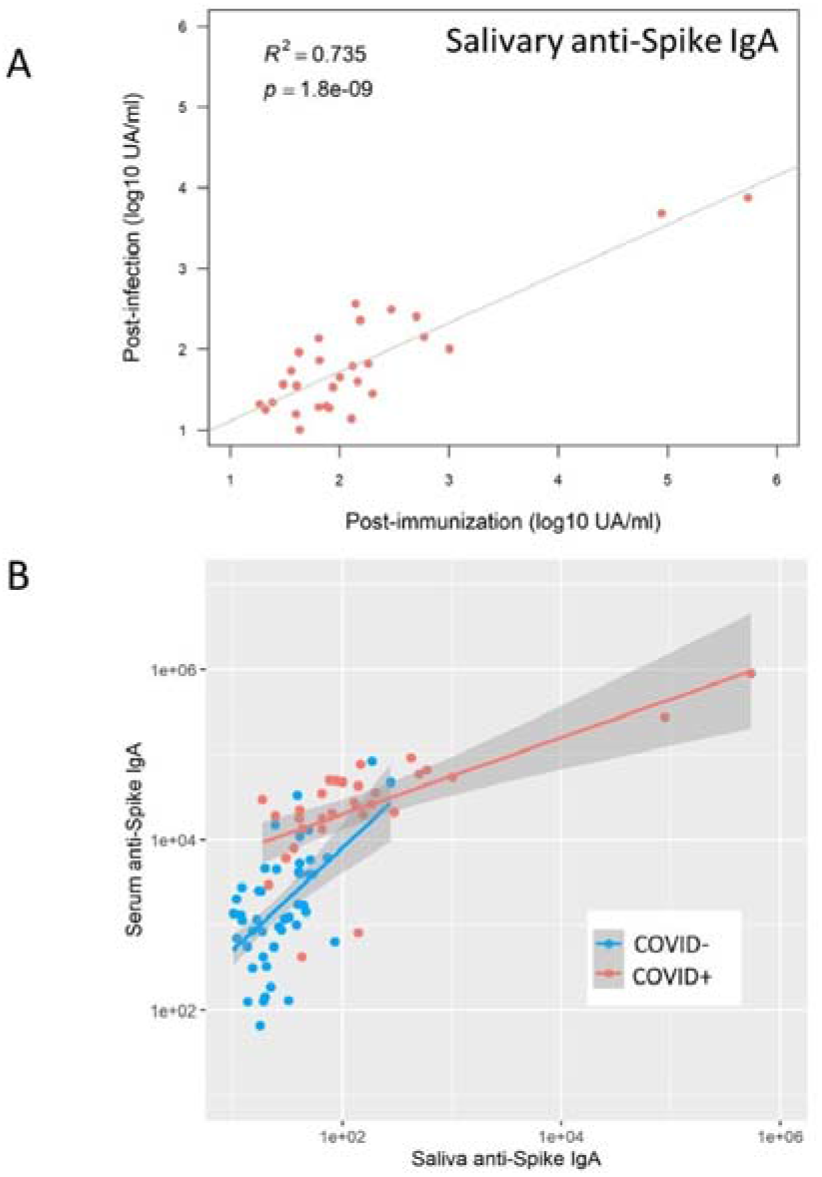
SARS-CoV-2 vaccination reactivated mucosal immunity in previously infected individuals. (A) Salivary anti-Spike IgA post-immunization (Visit 3) positively correlated with salivary anti-Spike IgA post-infection (Visit 1) in previously infected individuals (Pearson correlation, R^2^=0.735, p<0.001). (B) After immunization, the correlation between saliva and serum anti-Spike IgA was well differentiated in naive (COVID-, blue) and previously infected (COVID+, red) individuals, underlining the reactivation of mucosal immunity in previously infected individuals only: COVID+ group (Adj R2=0.44, slope = 0.45) ; COVID-group (Adj R2=0.31, slope = 1), p<0.001.

After vaccination, seroneutralization titers (Supplementary figure S8 and S3D) highly correlated with serum anti-Spike/NTD IgA level (Spearman, rho=0.92, p=0.0001), saliva anti-Spike/NTD IgA level (Spearman, rho=0.84, p=0.003), serum anti-Spike IgA (rho=0.86, p=0.0001) and with a lower correlation with serum anti-Spike/RBD IgA (Spearman, rho=0.36, p=0.005). The post-vaccine titers of anti-SARS-CoV-2 Spike IgA in serum and saliva did not modify the initial level of anti-NL63 and 229E hCoV Spike IgA signals (supplementary figure S9 and S10), but highly increased anti-SARS-CoV-1 Spike signal and slightly anti-OC43 and HKU1 anti-Spike signals (beta-coronaviruses) in serum, confirming that the correlation between human coronaviruses may interfere with the serodiagnosis shortly after vaccination.

Altogether, these results highly suggested that vaccination with mRNA and ChAd-vectored vaccines reactivated mucosal immunity in previously infected COVID-19 individuals but was not sufficient to induce an effective mucosal response in naive individuals.

### 2.5. Anti-SARS-CoV-2 Spike/NTD IgA titers induced by COVID-19 are significantly higher in individuals suffering from persistent smell or taste disorders one year after the acute infection

During the longitudinal follow-up of the cohort, the clinical status was evaluated at each visit. An important proportion of COVID-19 infected individuals had a significant persistence of symptoms after one year (43.5%), especially taste disorders, including dys-, hypo- and ageusia (n=38/102 in the cohort, n=10/58 in our study) and smell disorders, including dys-, hypo- and anosmia (n=42/102 in the cohort, n=13/58 in our study) (Supplementary Figure S11). We compared anti-SARS-CoV-2 IgA response in serum and saliva after infection from individuals with smell (Figure 8A) or taste disorders (Figure 8B) persisting less or more than one year after the acute infection with SARS-CoV2 ancestor Wuhan strain. We found out that patients with smell and taste disorders persisting more than one year had higher titers of anti-SARS-CoV-2 IgA in saliva at enrollment (V1), significantly targeting the Spike/NTD (taste disorders, p=0.02; taste disorders, p=0.04) but not in serum. As Spike-NTD specificity was positively correlated with higher seroneutralization titers, it suggests that it could be an IgA target of clinical relevance.

**Figure 8:**
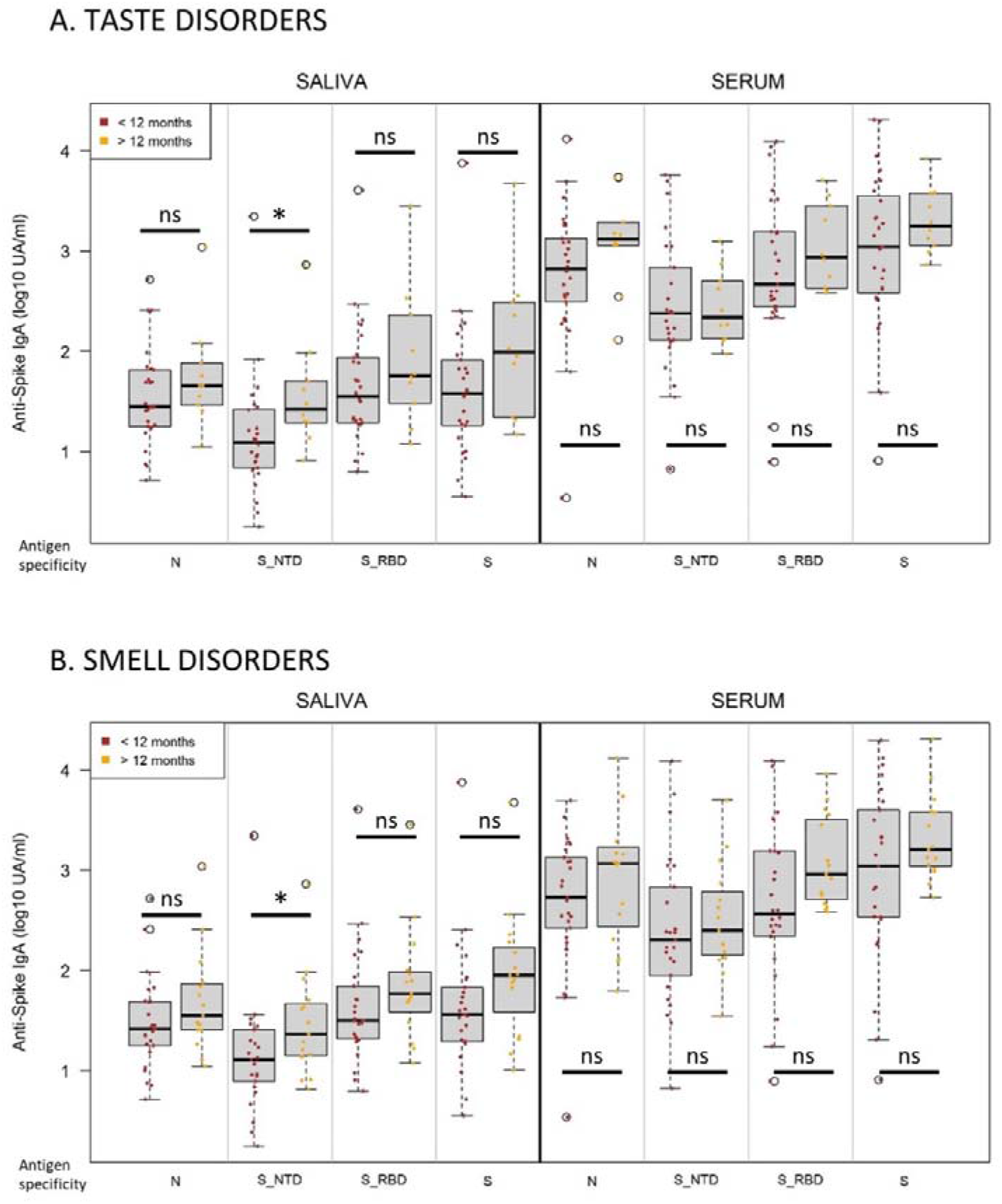
Persistent taste and smell disorders more than one year after infection are associated with higher initial titers of anti-Spike NTD IgA. (A) Normalized log-10 transformed serology titers of anti-SARS-CoV-2 IgA (log10 UA/ml) in saliva (left) and serum (right) measured at visit 1 by MSD technology from individuals suffering from taste disorders persistent for less (brown, n=28) or more (orange, n=10) than one year. (B) Normalized log-10 transformed serology titers of anti-SARS-CoV-2 IgA (log10 UA/ml) in saliva (left) and serum (right) measured at visit 1 by MSD technology from individuals suffering from smell disorders persistent for less (brown, n=27) or more (orange, n=15) than one year. Antigen specificity corresponds to the Nucleocapsid (N), Spike (S), Spike-receptor binding domain (S_RBD) and Spike-N-terminal domain (S_NTD). Kruskal-Wallis test: ns=not significant; * p<0.05.

## 3. Discussion

We followed SARS-CoV-2 IgA response in serum and saliva one year after a COVID-19 outbreak (SARS-CoV-2 Wuhan strain) in healthcare workers. By exploring its antigen specificity and kinetics, we aimed at better understanding the induction and persistence of mucosal immunity after infection and vaccination. We showed different kinetics and level of anti-Spike IgA in serum and saliva after infection, the first being sustainable up to 16 months after a first decline, the second being lost after 6 months. The antigen specificity of anti SARS-CoV-2 IgA unveiled the Spike/NTD as an important target, with positive correlations between serum anti-Spike/NTD IgA and titers of neutralizing antibodies, and saliva initial anti-Spike/NTD IgA levels and long-term persistence of smell and taste disorders. Vaccination with mRNA or Adenovirus-vectored vaccines reactivated a mucosal response in previously infected individuals but failed to induce a significant mucosal response in naive individuals, as already observed on a more limited cohort (25). As breakthrough infections have been correlated with IgA levels (16), IgA production may be a key point to address the issue of controlling the infection locally and preventing the transmission from vaccinated individuals. Most efficient vaccine platforms or route of administration inducing mucosal immunity are needed in the future for COVID-19 control, as already shown in preclinical studies (7, 9).

We used saliva IgA as a surrogate marker of mucosal immunity (10). Indeed, two major antibody classes operate in this fluid: secretory IgA and IgG. Most IgG in saliva are derived from serum whereas IgA are mostly synthesized by plasma cells in salivary glands, reflecting a local immune response that is generated independently from systemic IgA (12, 26). Saliva constitutes an interesting fluid offering an insight into the mucosal immune response (27), and a window into the plasma due to vascular leakage from the gingival crevicular epithelium (28, 29). Saliva is easier to sample and more accepted than nasal swab or nasal epithelial lining fluid. Previous studies on respiratory viruses, like influenza virus have already confirmed that saliva is good proxy at deciphering mucosal immunity (30-32).

The kinetics and antigen specificity of anti-SARS-CoV-2 IgA in our study underlined the sustainability of anti-Spike IgA response in serum over 16 months, after a first decline. This sustainability of the IgA response has already been described over 115 days post-infection with a rapid decline of IgA and IgM responses after a peak at days 16-30 (15), and for up to 8 months (33) and one year (34, 35). We were not able to catch the peak response in our cohort but we confirmed the decline between enrollment (early time post-onset of symptoms) and 6 months later, and the stability between 6 and 12 months. Early SARS-CoV-2 humoral response were shown to be dominated by IgA antibodies that contributed greatly to virus neutralization, in serum and saliva (36). By studying the antigen-specificity of the Spike response, we identified Spike/NTD as a specific IgA target that correlated with seroneutralization titers. A shift of the humoral response toward the Spike/NTD domain had previously been described after the second boost, despite the fact that some Omicron sublineages displayed several NTD mutations (37). Monoclonal antibodies targeting the Spike/NTD have shown in previous studies their ability to inhibit cell-to-cell fusion, to activate effector functions and to protect Syrian hamsters from SARS-CoV-2 challenge (38). Although, serum IgA titers have been correlated with the severity of the disease (39, 40). In our cohort of healthcare workers, the acute symptoms were mainly mild and could not been associated with the anti-Spike/NTD IgA titers. In saliva, IgA mainly targeted the Spike protein (including RBD and NTD specificity), with low titers constantly declining over the year of the follow-up. In previous studies, salivary IgG in mild-COVID-19 were detectable for up to 9 months post-recovery, whereas salivary IgA were short-lived. But salivary dimeric IgA were 15 times more potent than IgA monomers in a pseudoneutralization assay (41).

In our cohort, we observed slightly higher serum IgA titers against hCoV-229E and hCoV-0C43 than hCoV-NL63 and hCoV-HKU1. This description was similarly observed in another cohort using a bead-based multiplexed assay (42) and may reflect previous exposure in the life to these different circulating human coronaviruses (hCoVs). We noticed cross-reactivity inside hCoVs genera in serum, and more broadly between genera in saliva. Cross-reactive antibodies to hCoVs have been already reported in serum, against beta-hCoVs preferentially (43). The differences observed between correlations in serum and saliva were not surprising, as salivary IgA are generated locally and independently of systemic IgA. Mucosal immunity could favor the acquisition of a broader antigenic specificity than systemic immunity. We showed after infection that the level of anti-SARS-CoV-2 IgA response reached the range observed for the other hCoVs.

After vaccination, we confirmed that previously infected individuals had higher titers of IgA in serum than naive individuals and only naive people benefited from a booster effect of the second dose (44-46). Notably, even after two doses, IgA titers from naive individuals did not reach the level of response of previously infected individuals. In our cohort, only anti-Spike/NTD IgA specificity in serum was associated with neutralizing antibodies after infection. After vaccination, Spike, Spike/NTD and Spike/RBD positively correlated with neutralizing antibodies, as observed in other cohorts (45), but we also noticed a positive correlation with Spike/NTD in saliva. In previous studies, a detectable neutralizing activity in saliva observed 2 to 4 weeks post-vaccination was described low and transient with a rapid decline (16, 47). The ratio of saliva/serum IgA clearly showed two differential profiles according to previous COVID-19 status. Salivary IgA were not induced after vaccination in naive individuals. Conversely, salivary IgA were highly reactivated after vaccination in individuals previously exposed to SARS-CoV-2, and did not mirror serum IgA, suggesting that systemic vaccination could reactivate previously acquired mucosal immunity. This observation emphasized the importance of the first encounter of the antigen that imprints the immune response. Immune imprinting is a phenomenon whereby initial exposure to an antigen effectively primes lymphocyte memory to determine the eventual destination of an activated T cell, and the generation of local tissue-immunity, but limits the development of a different pattern of response that would be necessary to protect against a variant or another strain of the same pathogen after a secondary encounter of the antigen. It is known for viruses like influenza that the mode of initial exposure to a virus or a vaccine affects both the strength of the response and the breadth of the imprint (48, 49).

Lastly, we noticed that individuals suffering from persistent taste or smell disorders more than one year after a mild COVID-19 had higher initial titers of saliva anti-SARS-CoV-2 IgA targeting more specifically the Spike/NTD than individuals who recovered before one year. After vaccination, there was no more difference between the 2 groups. Persisting olfactory and gustatory dysfunctions were commonly reported, and probably underestimated (50). We could not correlate the level of the Spike/NTD-specific antibody response with the initial viral load as the outbreak took place very early, in March 2020, before semi-quantitative molecular diagnosis tools were deployed at our hospital. Studies in animals associated these symptoms with viral persistence and inflammation in the olfactory epithelium (51), but the underlying mechanisms of these persistent dysfunctions are still debated (19). Recent studies have started to explore the involvement of humoral responses in systemic (52, 53) and mucosal compartments (24). In accordance with their results, we did not observe a major difference in serum IgA regarding the duration of olfactory symptoms. Saussez et al. showed that persistent olfactory outcomes at 2 months were associated with lower saliva and nasal IgG1, without any modification of mucosal IgA (24). However, they did not investigate the level of specific anti-Spike/NTD IgA. Our observation was done on few subjects and needs to be confirmed in other cohorts. If confirmed, further studies will be needed to explore the underlying mechanism. It would also help physicians to predict the outcome of smell and taste disorders in order to address rapidly individuals at risk to otorhinolaryngology specialists.

## 4. Material and methods

### 4.1. Ethical statements

Ethical approval of the cohort study “Immuno-Covid Percy” was given by the Committee for Protection of Persons engaged in clinical research (CPP 20.05.25) and was registered in clinicaltrials.gov (NCT04408001). A written consent was obtained from all participants prior enrollment. The participants did not receive any compensation. Blood and saliva samples were anonymized, handled aliquoted and stored at appropriate temperature (−20°C for the saliva and the serum after blood centrifugation).

### 4.2. Cohort follow-up

The cohort follow-up started after a major outbreak among healthcare workers at the military teaching hospital Percy (HIA Percy) at the end of February 2020. The study population was constituted of hospital workers, including caregivers but also administrative staff. Subjects with a severe form of COVID-19 or immune disorders were excluded. At the initial visit (V1, from day 8 to day 118 after the onset of symptoms), volunteers were included, examined and followed at 6 months (V2, from day 193 to 305 post-onset of symptoms) and 12 months later (V3, from day 375 to day 479 post-onset of symptoms, ie. up to 16 months after infection). At each consultation, blood and saliva were sampled and the volunteers completed a medical form (Supplementary method). Convalescents (COVID+ group) and uninfected individuals (COVID-group) were defined on their SARS-CoV-2 genome detection (PCR) and/or total immunoglobulin serology (ELISA) against different SARS-CoV-2 Spike and Spike/RBD (Supplementary method).

### 4.3. Total immunoglobulin detection by Electrochemiluminescence assays

In order to confirm the serum status of each participant, total immunoglobulin reactivity of human sera samples was tested against SARS-CoV-2 N and RBD at each sampling timepoint (V1, V2, V3) using the Elecsys® electrochemiluminescence double-antigen sandwich assay performed on Cobas e601. The Elecsys® anti-SARS-CoV-2⍰Spike RBD is a quantitative immunoassay. According to the manufacturer, a corrective factor (1.029) was applied to convert the results obtained in units per mL to WHO International Standards for anti-SARS-CoV-2 antibodies in BAU/mL (Binding Arbitrary Units per mL). For undiluted sera, SARS-CoV-2 RBD antibodies measuring range covered from 0.4 BAU/mL to 257 BAU/mL and values higher than 0.8 BAU/mL were defined as positive. For sera whose values were below the detection limits, automatized dilutions (1/100e and 1/10 000e) provided by the supplier were used. The Elecsys® anti-SARS-CoV-2 Nucleocapsid assay provided a qualitative detection of antibodies using a recombinant nucleocapsid protein. The result of SARS-CoV-2 N antibodies was automatically computed in a cutoff index, with values ≥1.0 interpreted as reactive corresponding to a positive result. Since these immunoassays were not available at the beginning of the study, retrospective dosing on frozen serum aliquots (maximum thrice thawed) were performed. Independent serum samples were tested to confirm the lack of long-term effect of freezing (more than 6 months) on the measuring results of these immunoassays (data not shown).

### 4.4. Clinical samples selected for IgA detection

A total of 400 volunteers achieved the totality of the longitudinal follow-up, including 249 individuals (62.3 %) remaining uninfected and 151 individuals (37.8 %) infected by SARS-CoV-2 (Supplementary Figure S1). For IgA detection in serum and saliva, we selected 75 naive individuals and 58 previously-infected individuals, based on the certainty of the diagnosis at enrollment (Table 1), including PCR and ELISA/ECLIA serology, the absence of a new COVID-19 infection during the longitudinal follow-up, and the quality/quantity of saliva samples (compliance to one-hour fasting before sampling, including non-smoking) (Supplementary figure S1).

### 4.5. Virus production

Vero cells (ATCC CCL-81) were grown in DMEM medium (Gibco Cat. No. 31966021, ThermoFisher Scientific, Waltham, MA, USA). At confluence, the Vero cells were harvested and subsequently seeded at 1.5 × 105/mL in 96-well plates in order to reach confluence after 72 h at 37°C in 5% CO2. Serial dilutions of stock viruses were made in the infection medium [DMEM supplemented with 50 U/mL penicillin, 50 mg/mL streptomycin (Gibco Cat. No. 15070063, ThermoFisher Scientific) and TPCK-trypsin (Sigma Cat. No. T1426) at a final concentration of 1 µg/mL]. For the neutralization test, SARS-CoV-2 (BetaCoV/France/IDF0372/2020 strain) from clade 19A was isolated by the National Reference Centre (CNR) for Respiratory Viruses (Institut Pasteur), as described previously (54). The BetaCoV/France/IDF0372/2020 strain was supplied by CNR for Respiratory Viruses hosted by Institut Pasteur, headed by Pr. Sylvie van der Werf. The human sample from which the BetaCoV/France/IDF0372/2020 strain was isolated was provided by Dr. X. Lescure and Pr. Y. Yazdanpanah from Bichat Hospital, Paris. Moreover, the BetaCoV/France/IDF0372/2020 strain was supplied through the European Virus Archive goes Global (Evag) platform, a project that has received funding from the European Union’s Horizon 2020 research and innovation program under Grant No. 653316.

### 4.6. Virus titration

Briefly, 50 µL of 10-fold serial dilutions of SARS-CoV-2 were inoculated into eight replicate wells. The 96-well microplates were incubated at 37°C in a 5% CO2 atmosphere, and the cytopathic effect was checked 5 days after inoculation. The virus titer was calculated according to the method of Reed and Muench (55).

### 4.7. Seroneutralization assay

At each sampling timepoint (V1, V2, V3), the presence of neutralizing antibodies was sought by a seroneutralization assay performed on Vero cells using the Institut Pasteur SARS-CoV-2 reference BetaCoV/France/IDF0372/2020 strain, in a BSL3 facility. Neutralizing antibodies (Nab) tests were performed in flat-bottomed microtitre plates (96 wells), 3 days after seeding the Vero cells. Two-fold serial dilutions of inactivated sera, starting at a 1/40 dilution, were mixed with an equal volume of SARS-CoV-2 (100 TCID50/50 µL), and incubated for 1 h at 37°C. One hundred microlitres of the serum–virus mixture and 100 µL of infection medium were inoculated in each well (four wells per dilution), and the plates were incubated for 5 days at 37°C in 5% CO2. The cytopathic effect was checked 5 days after inoculation. Neutralization titers were expressed as the inverse of the final dilution of serum that neutralized 50% of the inoculated wells. As no cytopathic effects due to serum cytotoxicity were observed at a dilution of 1/40, a neutralization titer of 40 was considered as positive.

### 4.8. Quantification of serum and salivary IgA

At each sampling timepoint (V1, V2, V3), IgA reactivity of human serum and salivary samples was quantified against SARS-CoV-2 Spike, Spike/RBD, Spike/NTD and Nucleocapsid, and the Spike of other human coronaviruses (SARS-CoV-1, alpha HCOV-NL63, alpha HCOV-229E, beta HCOV-OC43 and beta HCOV-HKU1) on a MesoQuickPlex SQ120 (Mesoscale discovery – MSD) using the V-PLEX COVID-19 Coronavirus Panel 2 IgA kit (MSD, K15371U) and according to the manufacturer’s protocol. Briefly, after a blocking step, wells from a 96-well plate were washed and incubated during 2 hours at 22°C with diluted human serum samples (1: 5,000) or diluted human saliva samples (1:25). After three washing steps, bound antibodies were labelled with SULFO-TAG^™^ anti-human IgA antibody during 1 hour at 22°C. After three washing steps, read buffer was added and plates were read using MSD QuickPlex SQ120 electrochemiluminescent detection system. Samples out of range were up diluted and retested. Data were processed using MSD’s Discovery Workbench version 4.0 and quantification was reported in Arbitrary Units/mL (AU/mL) based on a reference standard curve included in the assay. To assess assay precision and inter-assay variability, three serological controls, containing known concentrations of human IgA against the panel targets’, were run in duplicate on each plate. The technical coefficient of variation (CV) of controls and the individual CV of negatives samples were calculated following the equation 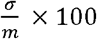 (*σ* = standard deviation of calculated IgA-concentration, m = mean of calculated IgA-concentration). The positive threshold of each test was determined by the quantitative variable maximizing both sensitivity and specificity (i.e. Youden index). The ROC curves are found in supplementary figure S2.

### 4.9. Statistical analysis

Statistical analyses were performed with R (version 4.2.5) using RStudio interface (version 2022.07.2+576). Graphical representations were made using ggplot2 package. Tests were performed in a bilateral way and p-values under 0.05 were considered statistically significant. A correction for multiple comparisons was applied using the Benjamini-Hochberg correction or Tukey HSD, resulting in a corrected p-value (false discovery rate) at or under 0.05. Nevertheless, we used a false discovery rate threshold of 0.2 for the association of persistent clinical symptoms with antigen-specific IgA titers in order to avoid missing potential important association due to the weaker number of subjects.

#### Discrete independent variables and quantitative dependent

Testing for group effect on a quantitative variable was performed with the following rank-based tests: Wilcoxon rank-sum test in unpaired two-group settings (comparing IgA concentration between naive and previously infected individuals); Kruskal-Wallis test with Dunn’s correction when comparing multiple unpaired groups (or two-way ANOVA when homoscedasticity and normality of residual was assessed); Friedman test with Nemenyi post Hoc test when comparing multiple paired groups (e.g. comparing concentrations at several timings) using the PMCMR plus package.

#### Quantitative variables

Linear mixed models were used to describe time effect on quantitative variables (lmer function, lme4 package). Following the Box Cox approach (powerTransform function, car package), concentrations were log10 transformed in these linear models to maximize linearity, homoscedasticity and normality of residuals. Simplification of complete models (free intercept and slope per individual for both time and period effects) to allow the model to converge led to models with free intercept per individual only. Slopes and intercepts were compared between conditions and periods using emmeans package. 95 percent profile confidence intervals for model estimates were extracted using the confint function, stats package.

## Data Availability

All data for this study are contained within the manuscript and the supplementary materials. The R script for running the analyses can be communicated on demand.

## 5. Informed Consent Statement

All participants gave their written consent to the study. Ethical approval was given by the Committee for Protection of Persons engaged in clinical research (CPP 20.05.25) and was registered in clinicaltrials.gov (NCT04408001).

## 6. Author Contributions

Conceptualization, AT, JNT, DR; methodology, JD, AG, LC, AF, HT, FJ, CH, EBD, MM, AT; software, JD, AT, MM; statistical analysis, JD, AT, MM; resources, AT, MM; data curation, JD, AT; writing—original draft preparation, JD, AT, MM; writing—review and editing, JD, AT, MM, JNT, EBD, AFR; visualization, JD, MM, AT; supervision, MM, AT; All authors have read and agreed to the published version of the manuscript.

## 8. Acknowledgments

The authors thank the healthcare workers from HIA Percy who massively enrolled in the cohort and gave their time for clinical research during the difficult period of the initial wave of the COVID-19 outbreak. The authors also thank the Percy ImmunoCovid group volunteers that were not directly involved in this work but largely helped to manage the cohort (Nassima Airapetian, Cyril Badaut, Marie Baruteau, Marie De Laage, Amir Dib, Corinne Jamet, Quentin Laborde, Carole Leclercq, Tiphaine Le Lièvre de la Morinière, Pierre-Yves Masse, Karine Michaud, Alice Nicolai, Amandine Steiger), all the team of the department of laboratory medicine who performed serological analysis (particularly Chrystelle Patole, Stéphanie Coppet, Pauline Chauvin) and PCR testing, and all the nurses of HIA Percy (especially from the aeromedical center, the neurophysiology and the anesthesia departments) who realized the blood samples. The authors are grateful to Mesoscale Discovery® for a free of charge loan of a Meso QuickPlex SQ 120 and to Catherine Drogou for her advice on saliva sample management. Finally, the authors address a special acknowledgment to Dr Thépenier for the statistical analyses on R software, its precious help to manage the cohort and for the critical revision of the manuscript.

