## Supplemental data for "Long-term systemic and mucosal SARS-CoV-2 IgA response and its association with persistent smell and taste disorders"

### Supplementary Figure S1

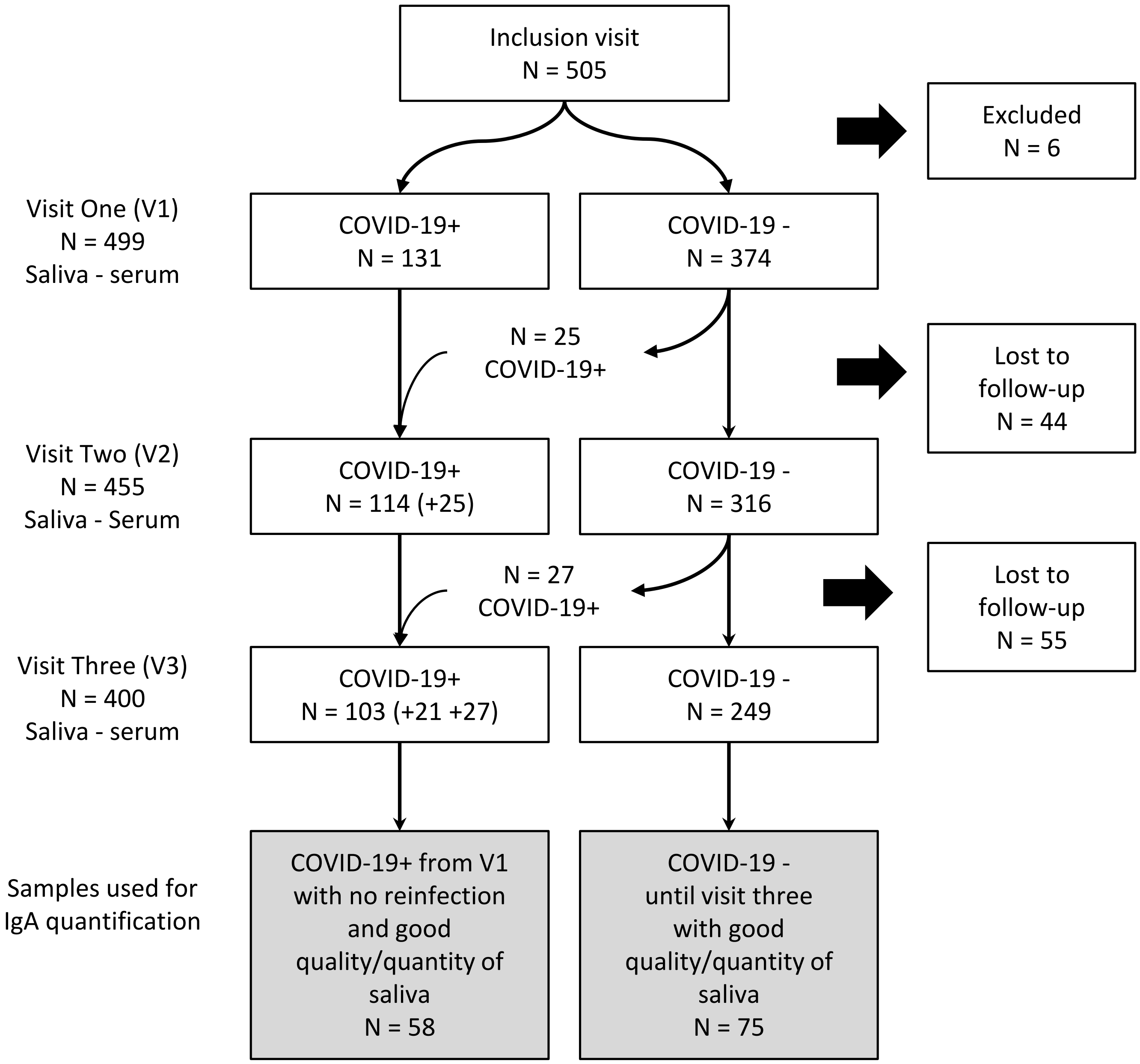


**Supplementary figure S1: Chart of the cohort follow-up and the number of selected samples for IgA quantification in sera and saliva for naive (COVID-19 -) and previously infected (COVID-19 +) groups.**

Supplementary method:

#### Cohort follow-up

At each consultation, blood and saliva were sampled and the volunteers completed a medical form. The initial medical form contained the following information: date of birth, gender, blood group, height, weight, smoking status, pregnancy, date of flu vaccination, medical and surgical histories, presence of any symptom consistent with a COVID-19 case and its time of onset, as well as the date and result of a related PCR if available (massive shortage at the beginning of 2020). At subsequent visits were notified the date of sampling, the weight, a new pregnancy, any clinical symptoms in favor of de novo or persistence of SARS-CoV-2 infection, any additional results of SARS-CoV-2 diagnosis (antigenic or PCR). The date and type of SARS-CoV-2 vaccination were noted since the vaccination campaign was initiated in early 2021 (few months before V3). Smell and taste disorders were assessed at each visit using a French adapted version of the validated Monell-Jefferson Taste-Smell Questionnaire (the Monell Chemical Senses Center).

#### SARS-CoV-2 genome detection by PCR:

The hospital policy during the COVID-19 early pandemic recommended to perform a PCR analysis for each symptomatic or case-contact worker (with a COVID-19 patient or co-worker). Several techniques were used by the hospital medical laboratory during the first wave including the TaqPath™ COVID-19 assay.

RBD/Spike indirect ELISA:

To address the lack of standardized serodiagnosis at the beginning of the pandemic, the initial COVID status of participants was determined by a classical indirect ELISA assay thanks to plasmids generously provided by Dr Florian Krammer (Amanat et al. Nat Med. 2020). Briefly, Sars-CoV2SARS-CoV-2-Spike and RBD antigens were produced in Expi293F cells (Life technologies #A14527) and purified on a HIS-Pur Ni-NTA resin (Thermofisher #88222). Plates were coated overnight at 4°C with 2 µg/ml of RBD or Spike. Sera were diluted at 1:200 for RBD and 1:500 Spike. The specific total IgG response was detected with a polyclonal anti-human IgG-HRP (Jackson ImmunoResearch #109-035-098). Positive sera against both targets (Spike and RBD) were assigned as COVID-19+ (Billon-Denis et al. Infection. 2020).

### Supplementary Table S1

| Biological fluid | Anti-SARS.CoV.2 IgA specificity | Loss of IgA | | | |
|---|---|---|---|---|---|
| | | Period 1 | Slope (mean[IC95]) in log(UA/mL)/day | Period 2 | Slope (mean[IC95]) in log(UA/mL)/day |
| **Serum** | | | | | |
| | Spike | V1-V2 | -1.28x10-3 [-2.35x10-3; -2.5x10-4] | V2-V3 | 1.75x10-4 [-9.68x10-4; 1.32x10-3] |
| | RBD | V1-V2 | -1.34x10-3 [-2.35x10-3; -3.12x10-4] | V2-V3 | 2.70x10-4 [-8.42x10-4; 1.42x10-3] |
| | NTD | V1-V2 | -1.86x10-3 [-2.85x10-3; -7.98x10-4] | V2-V3 | 2.93x10-4 [-8.01x10-4; 1.68x10-3] |
| | Nucleocapsid | V1-V2 | -3.01x10-3 [-3.89x10-3; -1.94x10-3] | V2-V3 | -1.73x10-4 [-1.49x10-3; 7.26x10-4] |
| **Saliva** | | | | | |
| | Spike | V1-V2 | -5.87x10-4 [-1.59x10-3; 6.1x10-4] | V2-V3 | -5.52x10-4 [-1.46x10-3; 9.06x10-4] |
| | RBD | V1-V2 | -4.68x10-4 [-1.32x10-3; 7.15x10-4] | V2-V3 | -5.56x10-4 [-1.2x10-3; 9.55x10-4] |
| | NTD | V1-V2 | -5.17x10-4 [-1.51x10-3; 4.52x10-4] | V2-V3 | -5.5x10-4 [-1.31x10-3; 7.86x10-4] |
| | Nucleocapsid | V1-V2 | -3.91x10-4 [-1.27x10-3; 4.2x10-4] | V2-V3 | -6.25x10-4 [-1.420x10-3; 3.31x10-4] |

**Supplementary table S1: mean slopes of IgA loss in serum and saliva between each visits of the cohort follow-up.**

### Supplementary Figure S2

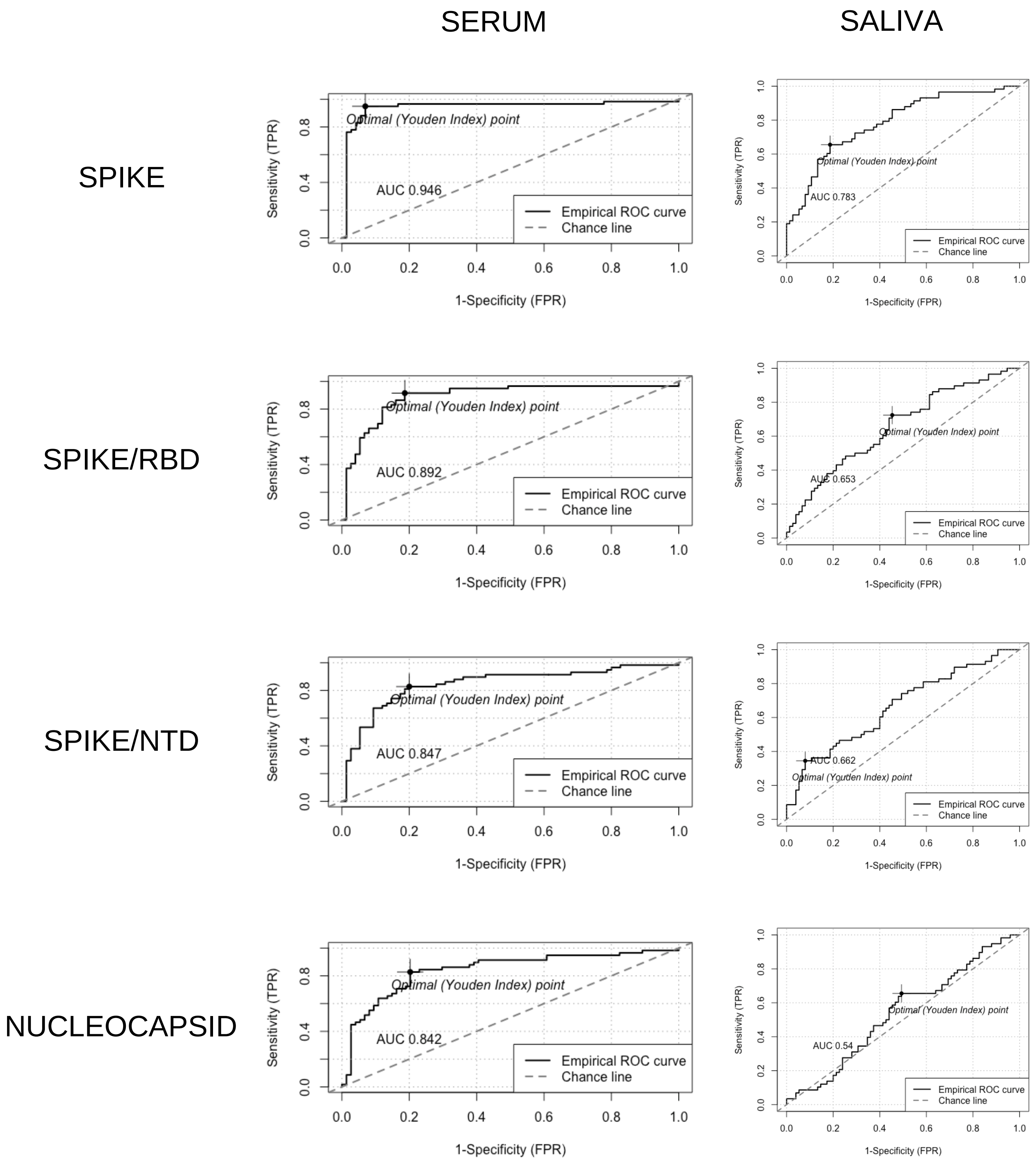


**Supplementary figure S2: ROC curves used to determine the IgA positivity threshold for each target (Spike, Spike/RBD, Spike/NTD, Nucleocapsid) in serum and saliva.**

### Supplementary Figure S3

#### A. Visit one (V1)

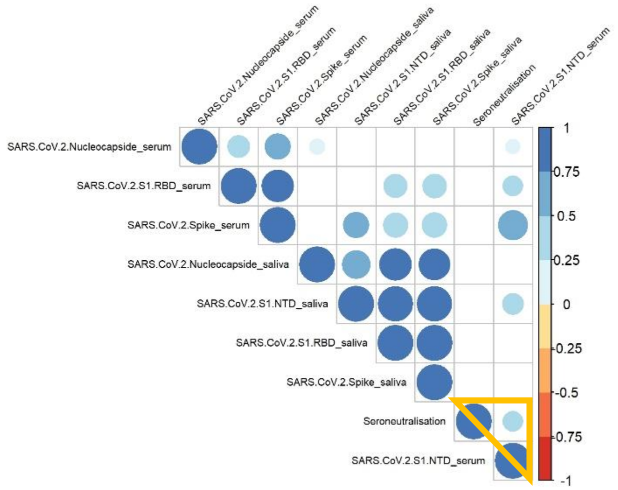


#### B. Visit two (V2)

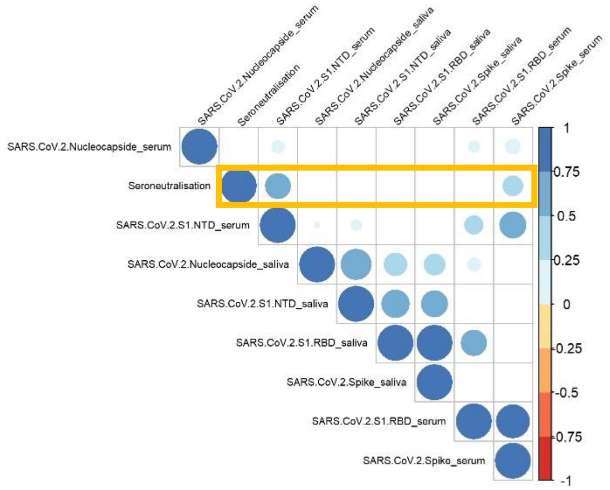


#### C. Visit three (no vaccine)

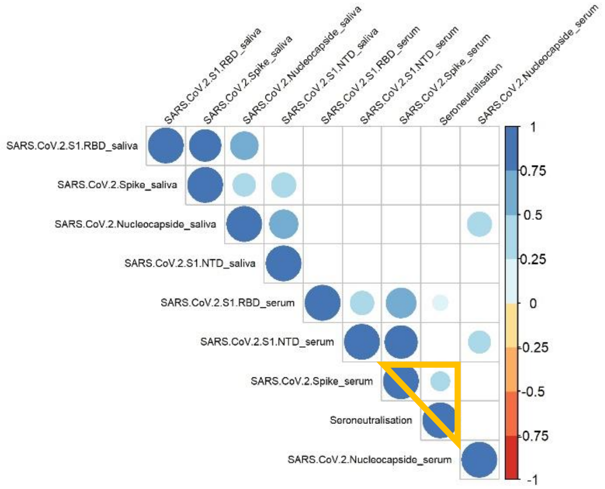


#### D. Visit three (after vaccine)

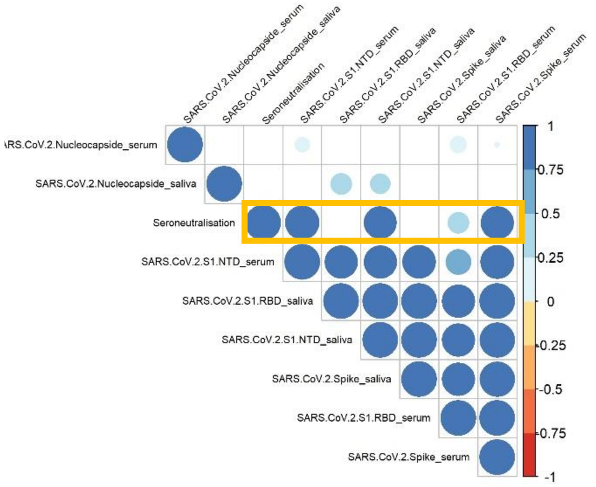


**Supplementary figure S3: Correlations of seroneutralization titers with SARS-CoV-2 antigen specificity targeted by IgA in saliva and serum.** Correlation matrices at V1 (A), V2 (B), and V3 without (C) or with (D) SARS-CoV-2 systemic immunization. Only significant correlations (Spearman, $p<0.05$) are represented on the matrices. The color and size of the dots (scale next to the graph) indicate the degree of correlation (Spearman, rho) between the different parameters (small to large indicating low to high correlation). The orange triangles or rectangles flank seroneutralization correlations at each visit.

### Supplementary Figure S4

A. COVID-19 – (serum)

B. COVID-19 + (serum)

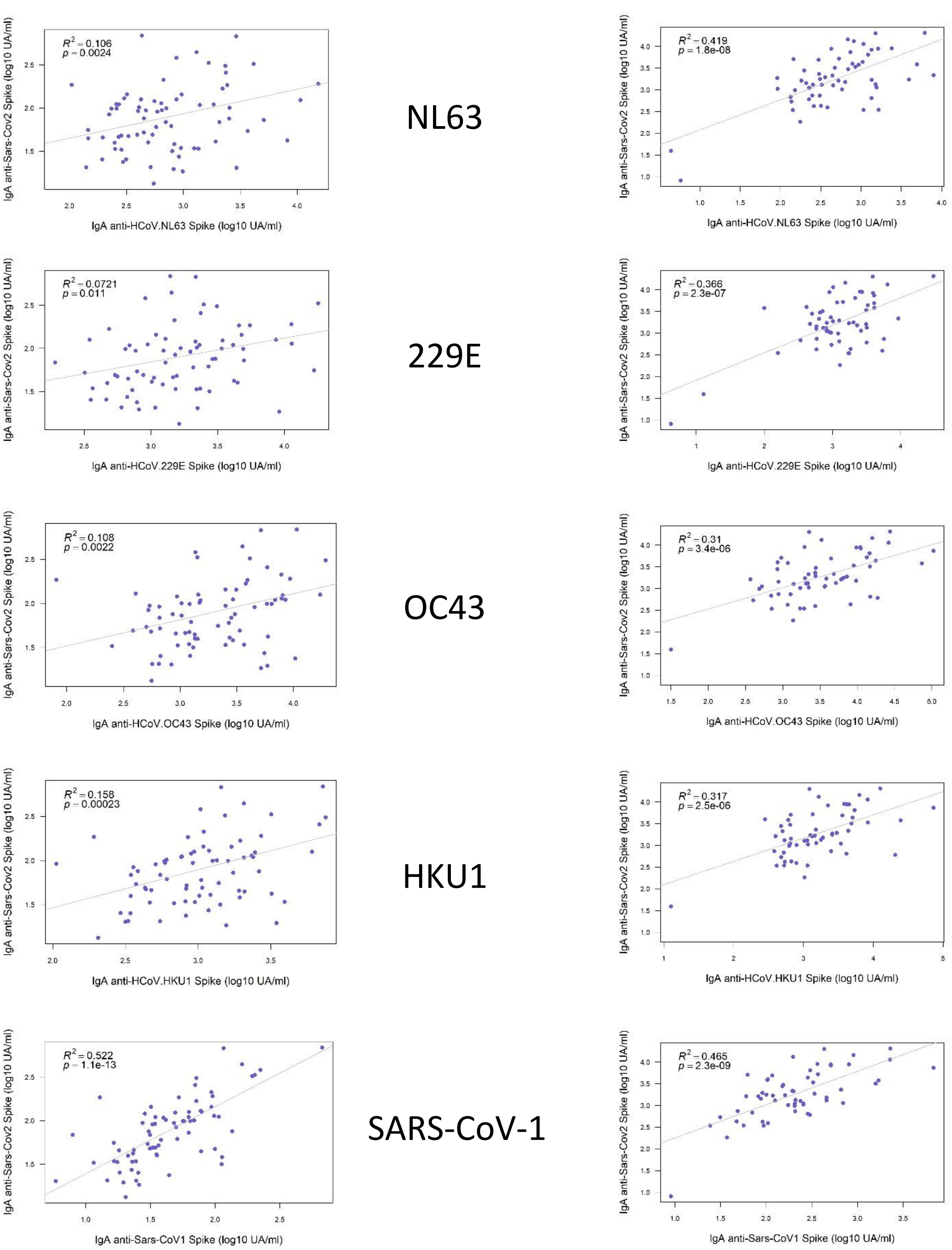


**Supplementary figure S4: Pearson correlation between anti-SARS-CoV-2 Spike IgA and anti-hCoV Spike IgA in serum.** (A) Naive individuals (COVID-19 -). (B) Previously infected individuals (COVID-19 +).

### Supplementary Figure S5

A. COVID-19 – (saliva)　　B. COVID-19+ (saliva)

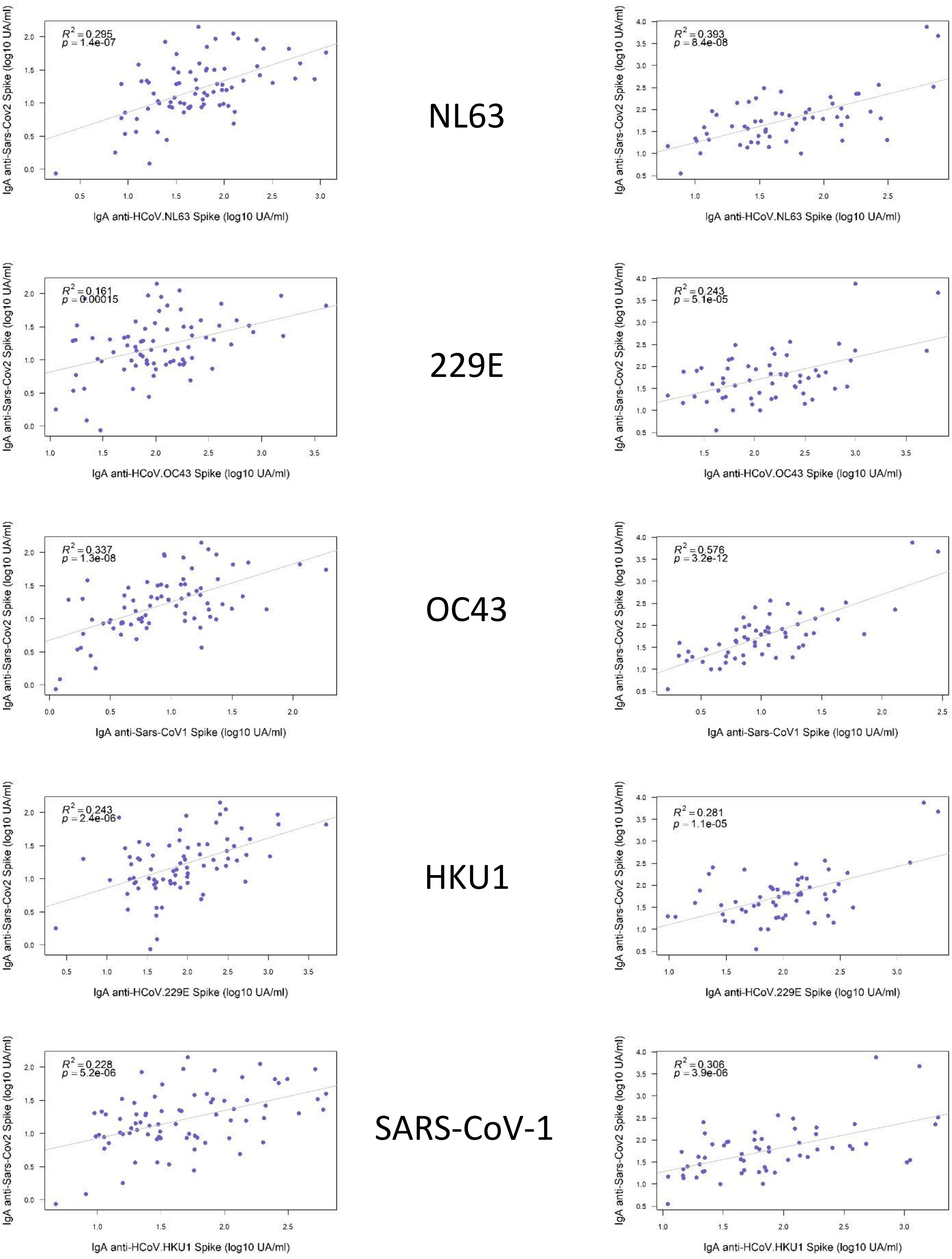


**Supplementary figure S5: Pearson correlation between anti-SARS-CoV-2 Spike IgA and anti-hCoV Spike IgA in saliva.** (A) Naive individuals (COVID-19 -). (B) Previously infected individuals (COVID-19 +).

### Supplementary Figure S6

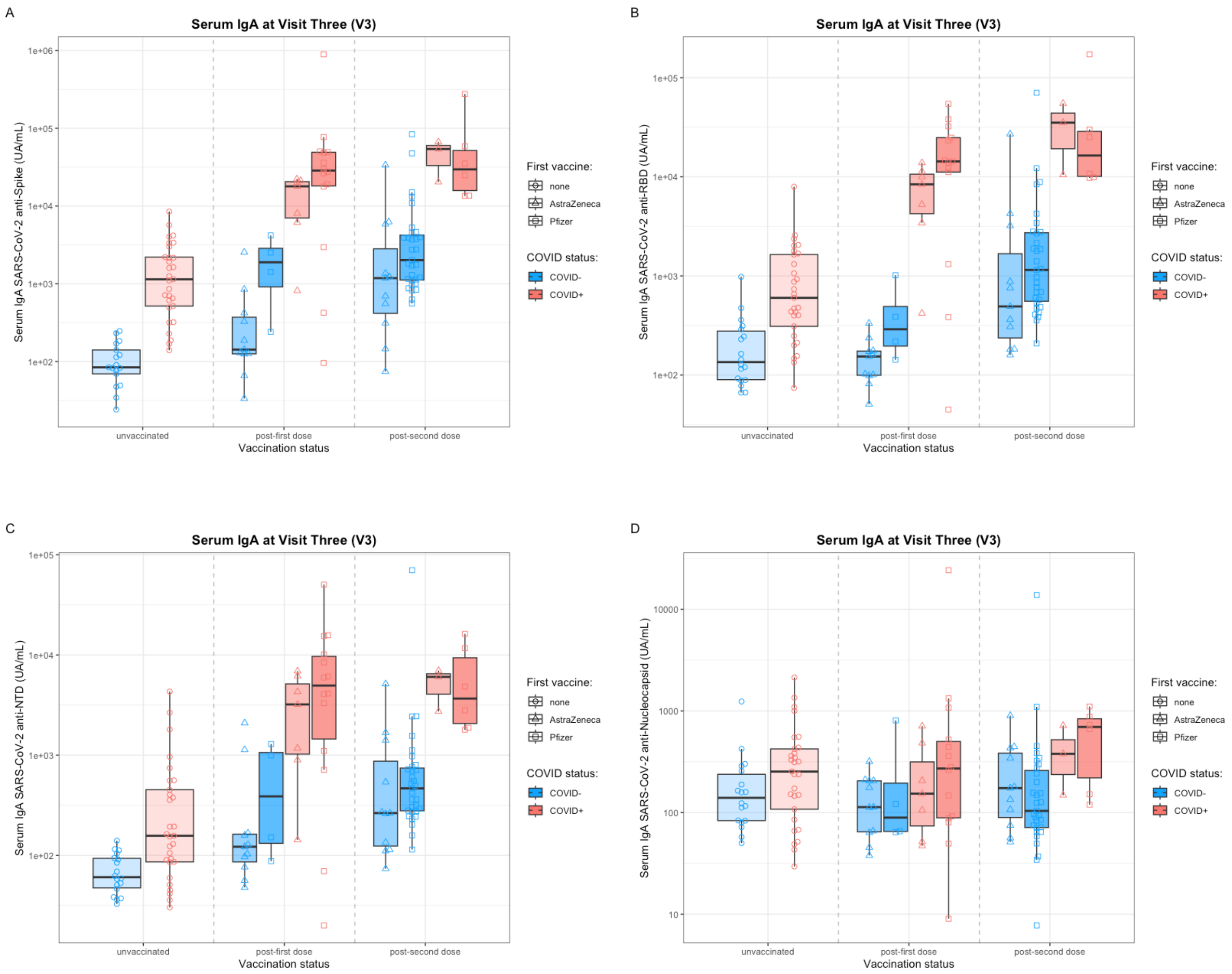


**Supplementary figure S6: Impact of the first vaccine (Astrazeneca vs Pfizer) on anti-SARS-CoV-2 IgA titers in serum in the presence (red) or absence (blue) of a previous COVID-19 infection.** (A) Serology IgA titers against the whole Spike. (B) Serology IgA titers against the Spike/RBD. (C) Serology IgA titers against the Spike/NTD. (D) Serology IgA titers against the Nucleocapsid.

### Supplementary Figure S7

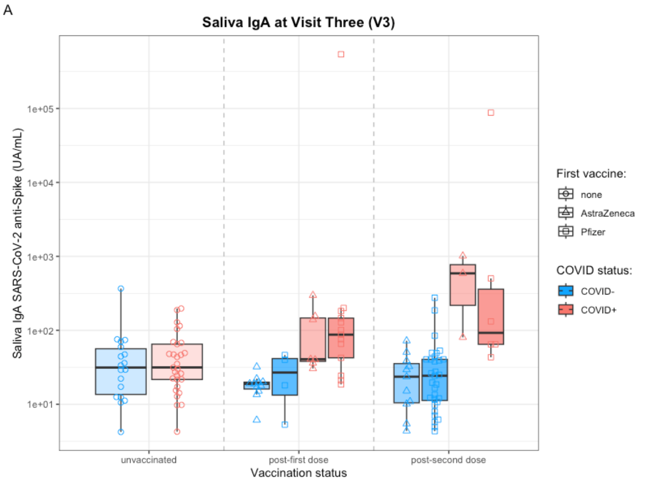


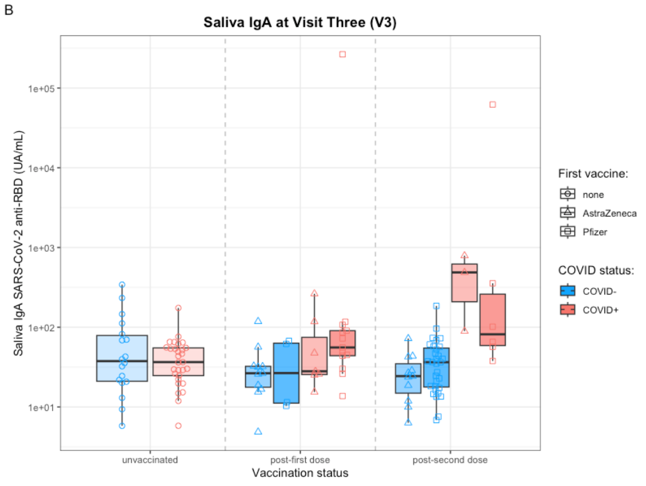


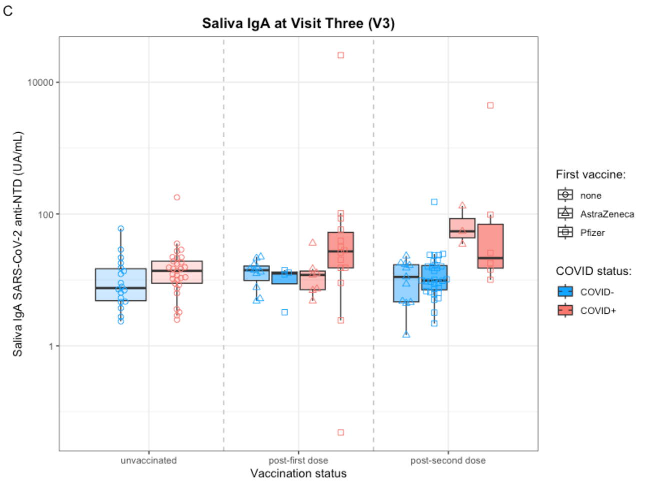


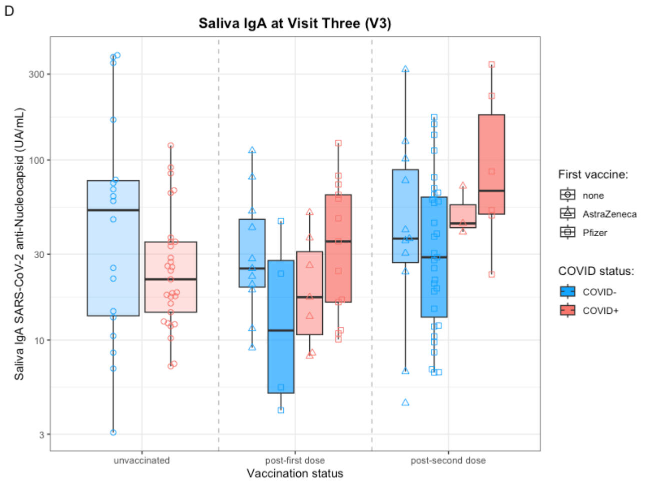


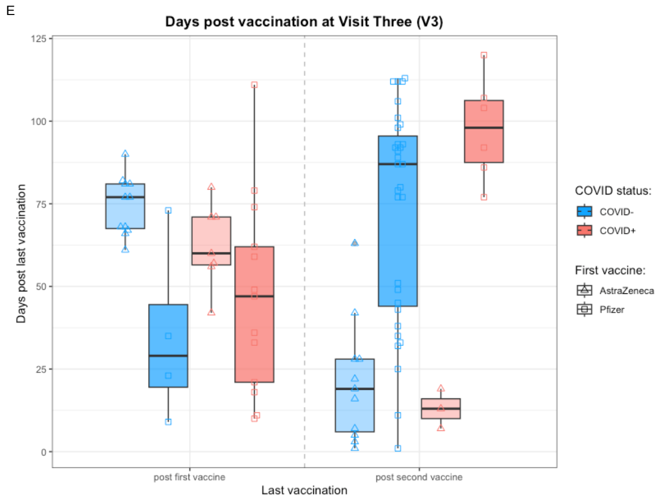


**Supplementary figure S7: Impact of the first vaccine (Astrazeneca vs Pfizer) on anti-SARS-CoV-2 IgA titers in saliva in the presence (red) or absence (blue) of a previous COVID-19 infection.** (A) Serology IgA titers against the whole Spike. (B) Serology IgA titers against the Spike/RBD. (C) Serology IgA titers against the Spike/NTD. (D) Serology IgA titers against the Nucleocapsid. (E) Days post-last vaccine dose at visit Three.

### Supplementary Figure S8

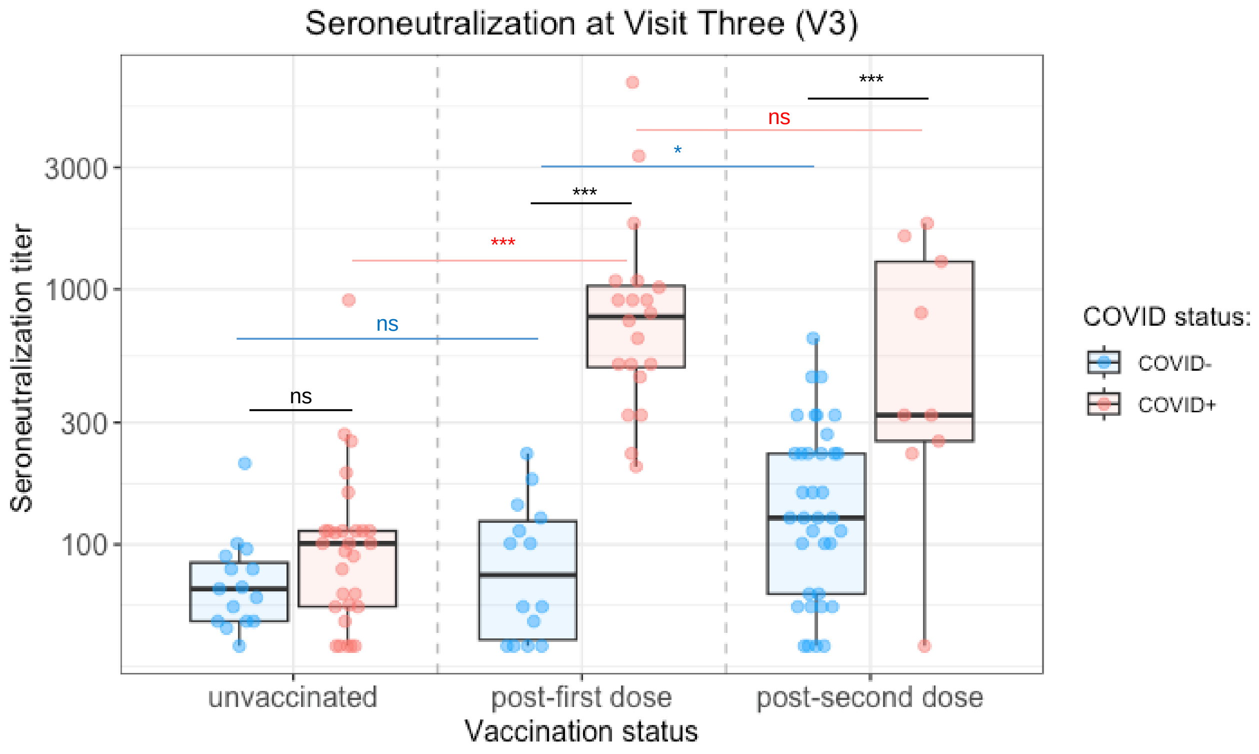


**Supplementary figure S8: Impact of the number of vaccine doses on seroneutralization titers against BetaCoV/France/IDF0372/2020 SARS-CoV-2 strain in the presence (red) or absence (blue) of a previous COVID-19 infection at V3.**
Two-Way ANOVA tests with Tukey post'hoc test: ns = not significant, * p<0.05 ; ** p<0.001; *** p<0.001.

### Supplementary Figure S9

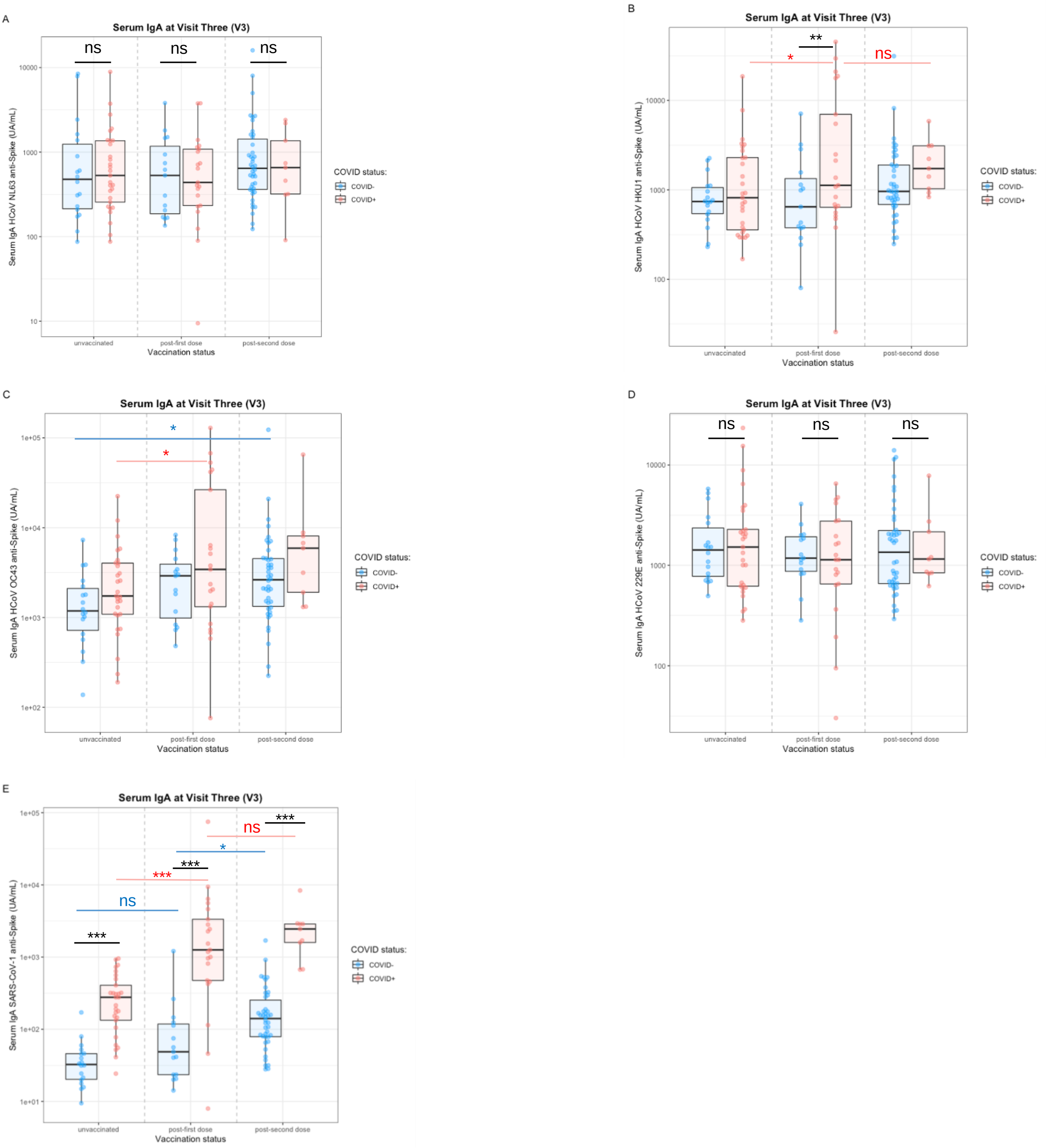


**Supplementary figure S9: Impact of the number of vaccine doses on other beta- and alpha-CoV anti-Spike IgA titers in serum in the presence (red) or absence (blue) of a previous COVID-19 infection.**

(A) Serology IgA titers against NL63-hCoV Spike. (B) Serology IgA titers against HKU1-hCoV Spike. (C) Serology IgA titers against OC43-hCoV Spike. (D) Serology IgA titers against 229E-hCoV Spike. (E) Serology IgA titers against SARS-CoV-1 Spike.

Two-Way ANOVA tests with Tukey post'hoc test: only significant differences are represented.

* p<0.05 ;  ** p<0.001; *** p<0.001.

### Supplementary Figure S10

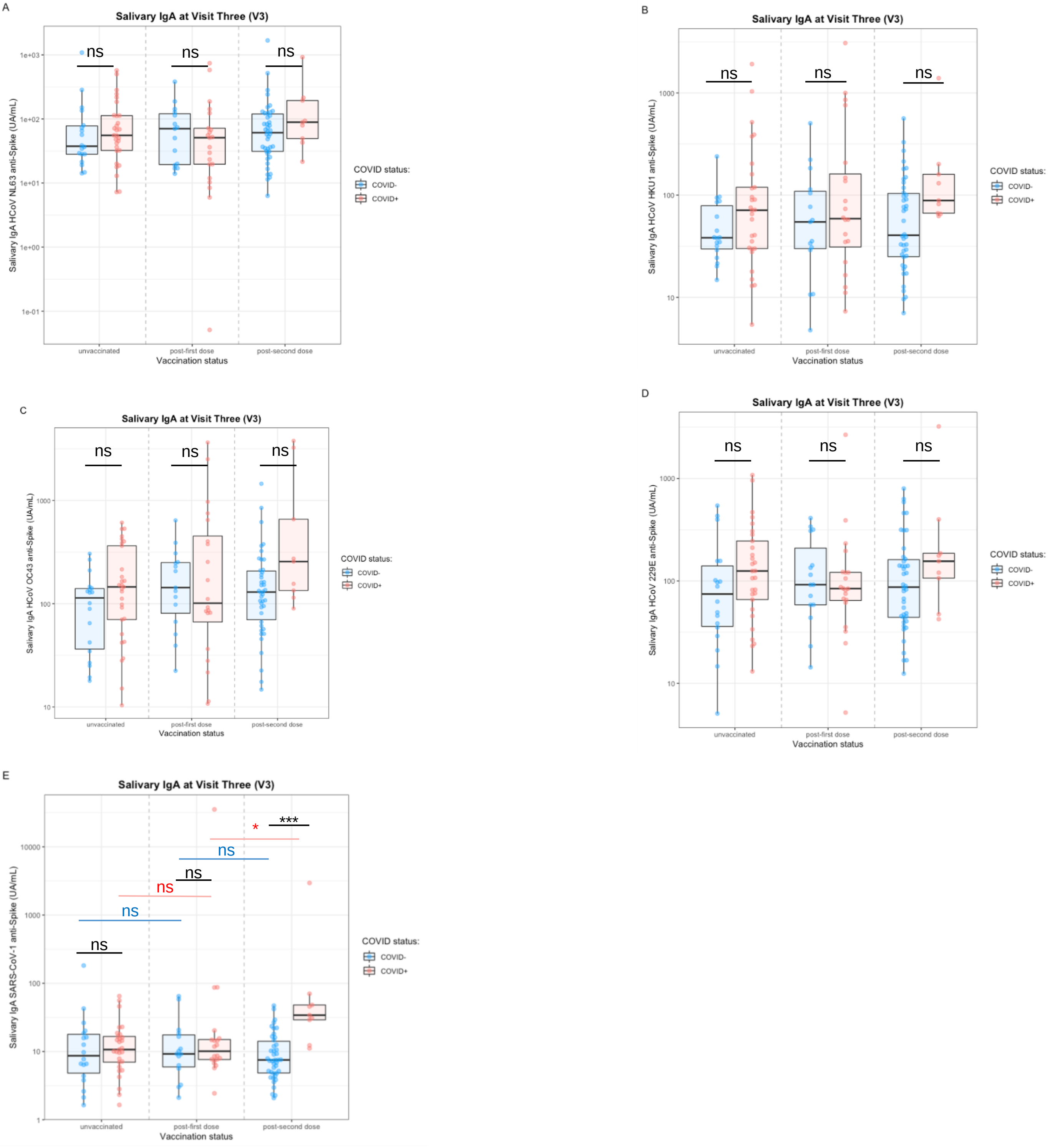


**Supplementary figure S10: Impact of the number of vaccine doses on other beta- and alpha-CoV anti-Spike IgA titers in saliva in the presence (red) or absence (blue) of a previous COVID-19 infection.**

(A) Salivary IgA titers against NL63-hCoV Spike. (B) Salivary IgA titers against HKU1-hCoV Spike. (C) Salivary IgA titers against OC43-hCoV Spike. (D) Salivary IgA titers against 229E-hCoV Spike. (E) Salivary IgA titers against SARS-CoV-1 Spike.

Two-Way ANOVA tests with Tukey post'hoc test: only significant differences are represented. * p<0.05 ; ** p<0.001; *** p<0.001.

### Supplementary Figure S11

A

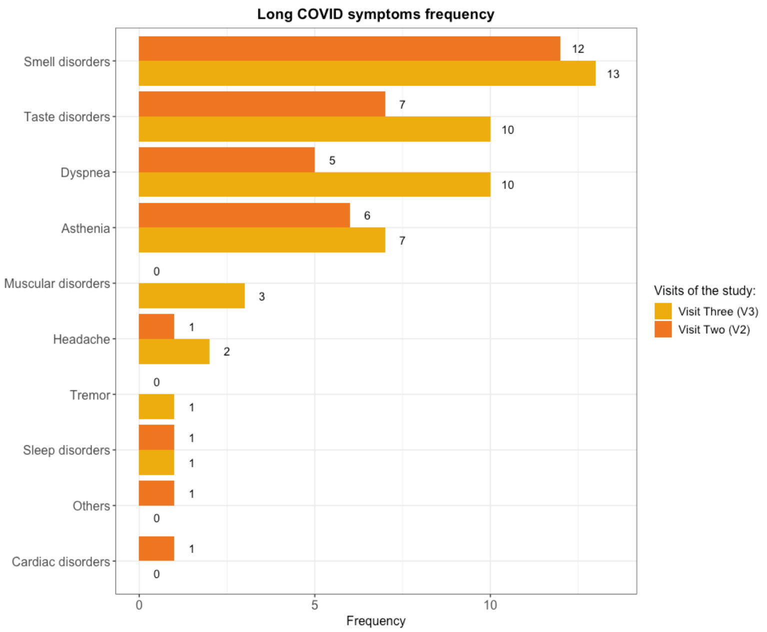


B

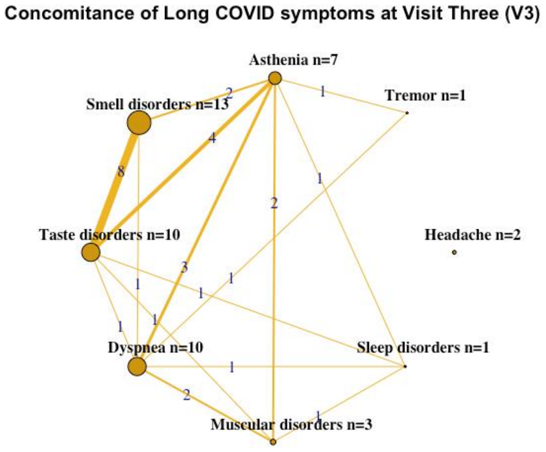


**Supplementary figure S11: Persistence of COVID-19 symptoms at visit 2 (V2) and visit 3 (V3, up to 16 months post-infection).**
(A) Frequency of persisting symptoms in previously infected individuals at V2 and V3. (B) Co-occuring symptoms from previously infected individuals at V3.
